# Mental Health Impact of COVID-19: A global study of risk and resilience factors

**DOI:** 10.1101/2020.05.05.20092023

**Authors:** Martyna Beata Płomecka, Susanna Gobbi, Rachael Neckels, Piotr Radziński, Beata Skórko, Samuel Lazzeri, Kristina Almazidou, Alisa Dedić, Asja Bakalović, Lejla Hrustić, Zainab Ashraf, Sarvin Es haghi, Luis Rodríguez-Pino, Verena Waller, Hafsa Jabeen, A. Beyza Alp, Mehdi A Behnam, Dana Shibli, Zofia Barańczuk-Turska, Zeeshan Haq, Salah U Qureshi, Adriana M. Strutt, Ali Jawaid

## Abstract

This study anonymously screened 13,332 individuals worldwide for psychological symptoms related to Corona virus disease 2019 (COVID-19) pandemic from March 29^th^ to April 14^th^, 2020. A total of n=12,817 responses were considered valid with responses from 12 featured countries and five WHO regions. Female gender, pre-existing psychiatric condition, and prior exposure to trauma were identified as notable risk factors, whereas optimism, ability to share concerns with family and friends like usual, positive prediction about COVID-19, and daily exercise predicted fewer psychological symptoms. These results could aid in dynamic optimization of mental health services during and following COVID-19 pandemic.

## MAIN TEXT

The emergence of novel severe acute respiratory syndrome coronavirus 2 (SARS-CoV-2) in December 2019 and the global spread of corona virus disease 2019 (COVID-19) has transpired as the most severe and publicized human crisis in recent history. As of April 19, 2020, the global burden of COVID-19 has exceeded 2.2 million cases and 152,551 deaths worldwide.^1^

Quarantine, isolation, and social distancing have been recommended by the World Health Organization (WHO), Center for Disease Control (CDC), and health officials worldwide to combat the spread of COVID-19.^2,3^ To adequately enforce implementation of these measures, one-third to half of the world is in complete lock-down as of April 19, 2020 ^4^ without a definitive end date determined. As a result of these extensive lock-down measures, economic markets have demonstrated alarming instability, with little indication of a timely recovery. The International Labour Organization recently reported that more than 700,000 jobs were lost in the past two months due to the COVID-19 outbreak and projected that up to 25 million jobs could be affected overall.^5^

The impact of COVID-19 on mental health of the masses has emerged as a matter of enormous concern.^6^ A number of factors related to COVID-19 can adversely affect the mental health of individuals, with an even higher risk in those predisposed to psychological conditions.^7^ Being in quarantine or isolation for extended periods of time has been associated with depression, anger, anxiety, and suicide as reported following the SARS epidemic of the early 2000s.^7^ Similarly, uncertainty of economic recovery and loss of job security are important factors previously associated with neuropsychiatric perturbations.^8-10^ Concerns have also been raised about increase in incidents of domestic violence and ‘screen time’ of individuals during the COVID-19 pandemic,^11-13^ which are known risk factors for the development or worsening of psychological conditions.^14^ Furthermore, fear and paranoia of being infected with SARS-CoV-2 and the stigma associated with manifesting symptoms such as cough or sneezing could negatively impact mental well-being.^15^ The fear of losing a loved one and the grief following loss are other potential disturbances to mental health accompanying disease outbreaks.^16,17^ Finally, it remains a consideration that SARS-CoV-2 may itself have neuropsychiatric manifestations as its effects on the nervous system are increasingly reported in patients who do not exhibit prominent respiratory tract symptoms.^18^

A number of studies from China have reported significant increases in symptoms of anxiety, distress, and risk of PTSD in students and health professionals assessed during the COVID-19 pandemic.^13,19-25^ A timely assessment on a global scale is paramount to display the mental health impact of the COVID-19 pandemic. With this data, health systems can strive to improve mental health services to reduce the long-term morbidity and mortality related to the COVID-19 crisis. Furthermore, this information could aid policymakers in improving the compliance of masses to the lock-down measures.^7^

To address this, we assembled a team of health professionals (neuroscientists, psychiatrists, psychologists, data scientists, and medical students) across all continents to develop a global study on the mental health impact of COVID-19. This study employs a fully anonymous online survey screening individuals in multiple countries for indicators and/or risk of general psychological disturbance, post-traumatic stress disorder (PTSD), depression, suicidal ideation, and concerns about physical health and appearance. The prevalence of these conditions was then cross-analyzed with participants’ demographics, opinions/outlooks, personality traits, current house-hold conditions, previous psychiatric disease history, and factors associated with COVID-19 to identify specific risk and resilience factors. We found alarming global trends for general psychological disturbances, risk for PTSD and depression, and suicidal ideation that were specifically predicted by participant demographics, personality traits, house-hold conditions, previous psychiatric disease and/or risk factor history and prediction about COVID-19 resolution.

## METHODS (ONLINE)

### Study Design

The study comprised a cross-sectional electronic survey-based assessment of individuals above the age of 18 years willing to participate in the study. The anonymous survey was conducted among participants from diverse demographic groups across continents using standardized self-report scales to screen for general psychological disturbance, risk for PTSD, and symptoms of depression. Specific responses were also independently assessed to screen for suicidal ideation and concerns for physical health and appearance. The survey was available online (placed on Google Forms platform) for a period of 15 consecutive days starting 18:00 Central European Time on March 29^th^, 2020 and concluding on 18:00 Central European Time on April 14^th^, 2020.

### Questionnaire development

The questionnaire was developed via close consultation between a neuroscientist, a neuropsychologist, a psychiatrist, a data scientist, and a psychiatry clinic manager. The questionnaire included closed-ended questions that assessed participant characteristics and opinions, and screened for neuropsychiatric conditions through standardized and validated self-report scales. The questionnaire prototype was prepared in English (Appendix 1) and translated into 10 additional languages (Arabic, Bosnian, French, German, Greek, Italian, Persian, Polish, Spanish, and Turkish; Appendix 2). The translation was performed by bilingual native speakers and vetted by volunteers native to those countries. The feasibility of each questionnaire was confirmed using pilot studies comprised of 10 participants each. These responses were excluded from the final analysis.

The questionnaires (Appendix 1) included a section on participant demographics (age, gender, country, residential setting, educational status, current employment status) house-hold conditions (working/studying from home, home isolation conditions, pet ownership, level of social contact, social media usage, time spent exercising), COVID-19 related factors (knowing a co-worker, friend, or family member who tested positive for or demised due to COVID-19, prediction about pandemic resolution), personality traits (level of optimism, level of extroversion), previous history of psychiatric disease and/or trauma, previous exposure to human crisis, and level of satisfaction with actions of the state and employer during the current crisis. All questionnaires were rated on binary (yes/no) responses or Likert-type scales.

The other sections contained general health assessment based on WHO Self-Reporting Questionnaire-20 (SRQ), Impact of Event Scale (IES), and Beck’s Depression Inventory II (BDI).^24,26,27^ These scales were chosen based on their common usage and efficacy in previously employed works studying the psychological impact of human crises, including the SARS epidemic.^28-36^ IES was purposefully adjusted to assess the impact of an ongoing event rather than a past event. For this purpose, the past tense was converted into the present tense in each question without changing the subject matter. This adjustment was performed in consultation with an independent neuropsychologist not involved in the study. For all scales, participants were prompted to think of and report their physical and psychological state during the preceding week.

### Ethical Considerations

Informed consent was obtained from each participant to allow anonymous recording, analysis, and publication of their answers. The data was collected in a completely anonymous fashion without recording any personal identifiers. This strategy ensured that the confidentiality of the participants was maintained throughout all phases of the study. The study procedures were reviewed and approved by University of Zurich Research Office for Scientific Integrity and Cantonal Ethics Commission for the canton of Zurich (Switzerland; Appendix 3), Nencki Institute of Experimental Biology, Warsaw (Poland; Appendix 4), Faculty of Medicine, University of Tuzla, Tuzla (Bosnia and Herzegovina; Appendix 5), and the executive board of the European MD-PhD association (EMPA).

### Data Collection

Using a non-randomized referral sampling (snowball sampling) method, participants were contacted by a team of 70 members (study authors and volunteers that have been acknowledged in the acknowledgement section) using electronic communication channels including posts on social media platforms, direct digital messaging, and personal and professional email lists. A concerted effort was made to ensure maximum participation from the countries that had the highest number of cases and >100 daily new cases (as reported on www.worldometers.info) as of March 29^th^, 2020. These countries included USA, Spain, Italy, France, Germany, UK, Iran, Turkey, and Switzerland. China was not included in this list (from here on referred to as the featured countries) due to the number of daily new cases being <100 during the data collection period. In addition to the most severely affected countries, a concerted effort was made to include two countries each from the 11^th^-20^th^ (Canada and Poland), and a country each from the 20^th^-30^th^ (Bosnia and Herzegovina), and 30^th^-40^th^ (Pakistan) most affected countries in the featured list. For the featured countries, national coordinators also reached out to at least 10 social media influencers and requested their voluntary help with the diffusion of the survey. The overall number of responses obtained via social media influencers was primarily a reflection of those from Pakistan, Spain, Switzerland, and USA. The data collection procedures were repeated at least thrice during the data collection period (March 29^th^- April 14^th^, 2020) with the aim to ensure participation of at least 250 participants each from the list of featured countries. This aim was achieved in all the featured countries with the exception of the UK, which was subsequently excluded from the featured list. For the non-featured countries, where the number of responses was less than 250 per country, the responses were grouped together based on the WHO regions (African Region AFRO, Region of the Americas PAHO, South East Asia Region SEARO, European Region EURO, East Mediterranean Region EMRO, and Western Pacific Region WMRO). WHO AFRO region was excluded from the analysis due to total number of responses being less than 250.

The data was collected exclusively online for participants under 60 years of age. For participants who were 60 or above, a special provision was allowed for assistance in recording their responses online as older adults are often not comfortable with virtual platforms.^37^

Our data collection strategy resulted in a total of 13,332 responses. Surveys completed by participants who were younger than 18 (n=34), those with missing responses for all dependent variables (n=112), filled the second time (n=325), missing geographic location (n=20), and from WHO AFRO region (n=24) were excluded from the final analysis. When the responses were missing for individual items, the missing data were considered null and excluded from the analysis for that particular variable. The number of participants for each of the featured countries and the regions encompassing the non-featured countries is represented in the Supplementary item S1.

**Item S1.**
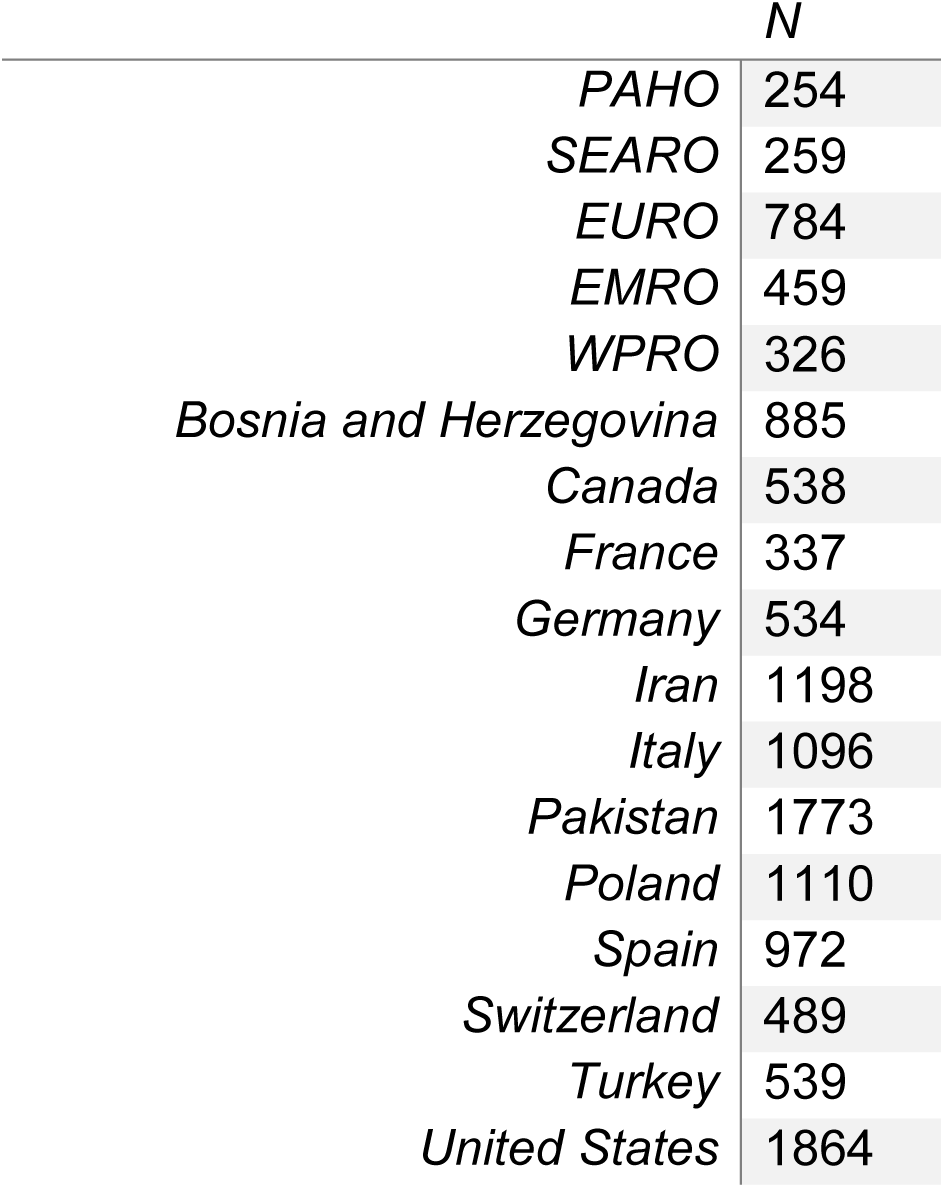
Number of participants per country and WHO region.

The snow-ball sampling method precludes us from inferring the response rate of the study. However, a minimum of 250 participants from the featured countries and the WHO regions with the non-featured countries was chosen to ensure that any within-group differences could be elucidated with a reasonable statistical confidence and an error margin <5%.

### Statistical Analysis

All statistical analyses were performed using R version v.3.6.3 and *Rstudio* (Rstudio team, 2015). All figures were produced using the packages *ggplot2* (Wickham et al., 2016) and *CGPfunctions* (Powell, 2020).

#### Non-adjusted analysis for SRQ, IES, and BDI scores

Mean scores with standard deviations were calculated for SRQ, IES and BDI scores from all valid responses (n=12,817) and compared across all of the following categorical predictors via Kruskal-Wallis tests with the Chi-square function. The categorical predictors included gender, residential status, education level, employment status, being a medical professional, working remotely from home, satisfaction with employer, satisfaction with the state (government), home-isolation status, interaction with family and friends, social media usage, ability to share concerns with a mental health professional, ability to share concerns with family and friends, prior exposure to a human crisis situation, previous exposure to trauma, level of extroversion, prediction about COVID-19 resolution and one’s self-determined role in the pandemic.

#### Multiple Regression Models for SRQ, IES, and BDI

Multiple linear and logistic regression models were built for SRQ, IES, and BDI using mean scores and cut-offs for respective categorical classification.

For linear regression, generalized linear models with the *glm function* were devised using the *lme4 package* (Bates et al., 2015). The three univariate linear regression models, one each for SRQ, IES, and BDI, were fitted and corrected for multiple comparisons followed by *glm function* analyses. Following the Bonferroni correction for multiple comparisons, the p-value threshold was set to 0.017. For each linear regression model, ‘age’ was entered as a continuous independent predictor whereas all aforementioned predictors were entered as categorical fixed effects. Poisson family and log link function were used to model BDI and SRQ factors. In order to choose the best model (based on Akaike information criterion; AIC or Bayesian information criterion; BIC) from the set of predictors, stepwise model selection was performed from the *MASS* package (Venables et al., 2002).

Logistic regression was performed to generate odds ratios (ORs) for SRQ, IES, and BDI using the following categorization scheme; SRQ: 0 = normal (0-7 points), 1 = concern for general psychological disturbance (8-20 points); IES: 0 = normal (0-23 points), 1 = PTSD is a clinical concern (24-32 points), 2 = threshold for a probable PTSD diagnosis (33-36 points), 3 = Severe condition (high enough to induce immunosuppression) (≥37 points). For generating ORs, the variables were regrouped as 0 = no concern versus any type of concern (1/2/3); BDI: 0 = These ups and downs are considered normal (1-10 points). 1 = Mild mood disturbance (11-16 points), 2 = Borderline clinical depression. (17-20 points), 3 = Moderate Depression (21-30 points), 4 = Severe Depression (31-40 points), 5 = Extreme Depression (>40 points). For generating ORs, the variables were regrouped as 0 = no concern versus any type of concern (levels 1/2/3/4/5). Cut-offs for SRQ, IES, and BDI were defined using least stringent thresholds for each of these measures from previous literature to ensure high sensitivity of the screening.^24-28^

Furthermore, three separated OR analyses were performed for suicidal ideation, and concerns about physical health and appearance based on relevant questions from BDI. For these models, reference level was set to 0= absence of symptom that was compared to presence of symptom (varying severity levels of the symptom regrouped into one category).

Finally, correlations between SRQ, IES, and BDI were performed through Pearson’s correlation test and illustrated as x~y plots. All statistical analyses were performed by the analysis team comprising MP, SG, PR, and AJ in consultation with ZB.

## RESULTS

### Regional and Worldwide Prevalence of Psychological Symptoms

A total of 12,817 valid responses were divided across USA (1864), Iran (1198), Pakistan (1173), Poland (1110), Italy (1096), Spain (972), Bosnia and Herzegovina (885), Turkey (539), Canada (538), Germany (534), Switzerland (489) and France (337). The remaining countries were grouped according to WHO regions, i.e. European region EURO (784), East Mediterranean region EMRO (459), Western Pacific region WPRO (326), South East Asian region SEARO (259), and region of the Americas PAHO (254). Significant (p<0.05) regional differences were observed for the psychological impact of COVID-19 (Fig. 1), with higher SRQ scores (indicating general psychological disturbance) in Bosnia and Herzegovina, Canada, Pakistan, and USA; and higher IES (indicating risk of PTSD) and BDI (indicating risk of depression) scores in Canada, Pakistan, and USA (Supplementary Item S2). Furthermore, higher prevalence of suicidal ideation was noted for Canada, Pakistan, and Poland (Supplementary Item S3)

**Item S2.**
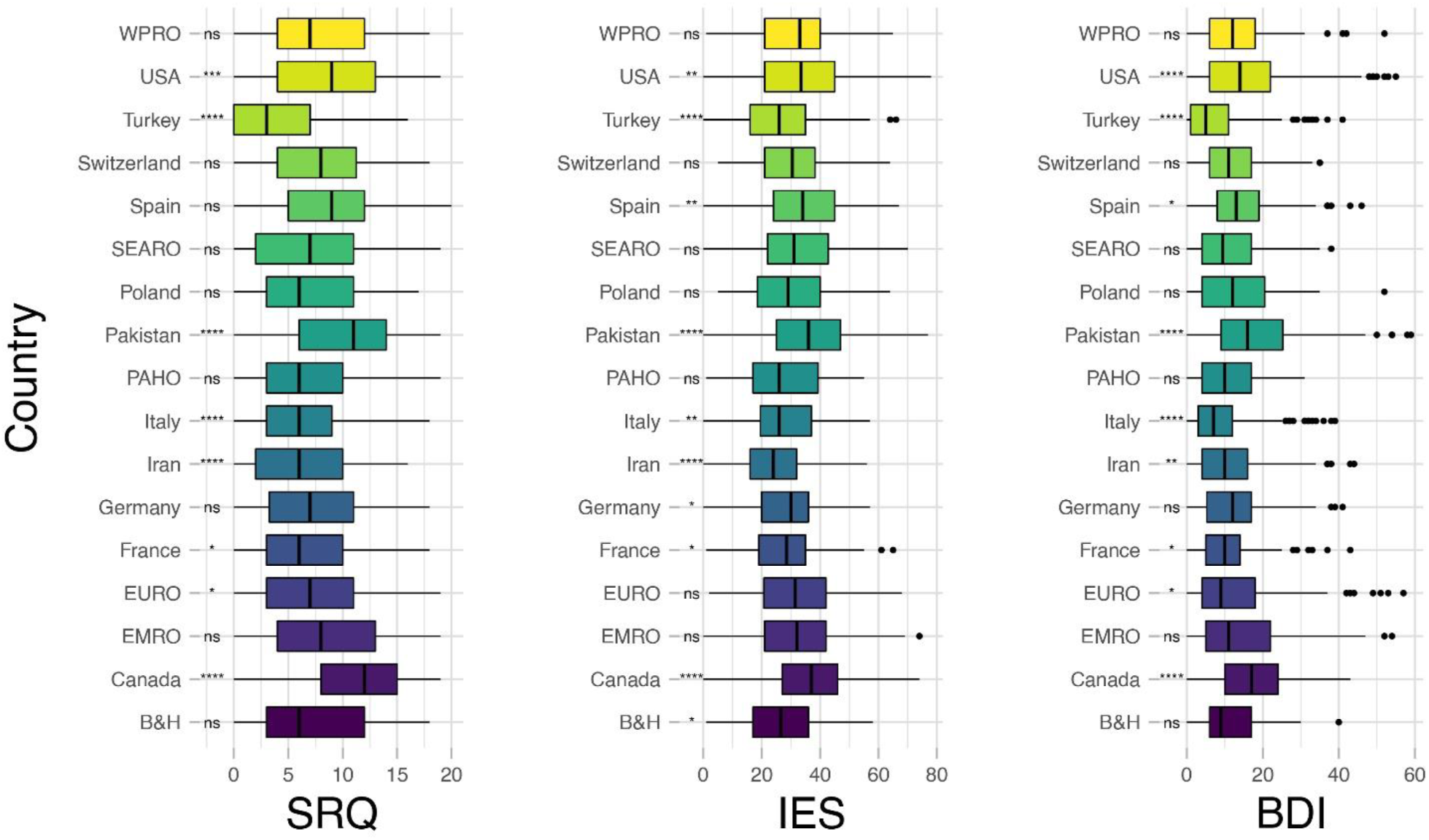
Differences across countries for SRQ, IES, and BDI scores. The boxplots show the distribution of scores for each country with the visualization of five summary statistics (minimum, maximum, median, first quartile, third quartile), and all outliers individually. Countries with significantly different scores are indicated on the left-hand side. The results were obtained performing pairwise comparisons with a non-parametric Wilcox test. **** p< 0.0001, *** p < 0.001, ** p < 0.01, * p < 0.05, ns= non-significant, B&H= Bosnia and Herzegovina.

**Item S3.**
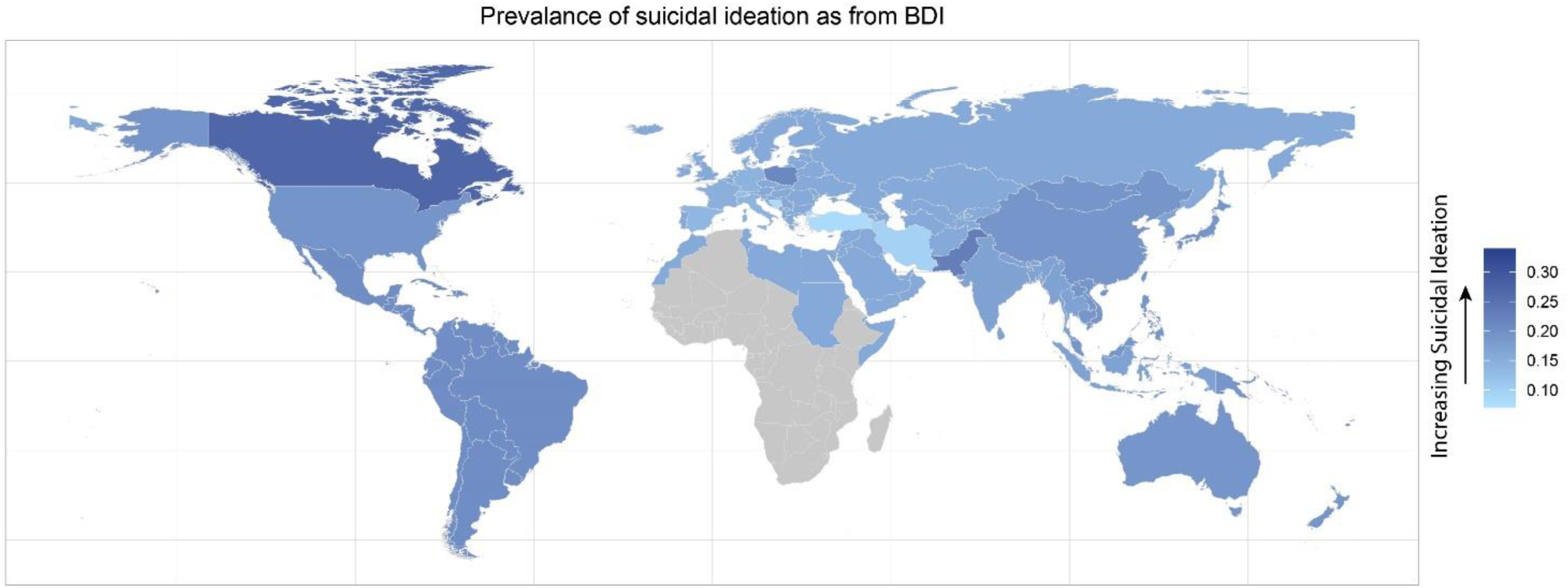
Geodemographic representation of Suicidal Ideation. The map presents mean scores in the question from BDI assessing suicidal ideation (scale 0-3). For this illustration, varying levels of suicidal ideation (1-3) are grouped together as 1, i.e., 0= no suicidal ideation, whereas 1 = any suicidal ideation. The mean scores are calculated separately for each of the featured countries, and for each of WHO regions using this binary classification for suicidal ideation.

There was a slight disproportion in valid responses, with higher numbers from those participants who were female (72.36%), residing in urban areas (82.87%), with advanced educational qualification, i.e., bachelor’s degree or higher (75%), working/studying remotely from home (64.4%), and currently under home-isolation with a partner/family (83.06%). Also of notable prevalence were factors, such as expressing satisfaction with COVID-19-related employer response (33.91%), being somewhat satisfied with COVID-19-related state response (37.08%), and spending less than 15 minutes on daily physical exercise (48.99%). A majority of participants also reported increased social media usage (65.15%), less-than-usual or minimal interaction with family and friends (70%), and feeling a sense of control in protecting themselves and others during the COVID-19 pandemic (80.86%). Details of participant demographics, household conditions, history of psychiatric conditions and exposure to trauma/crisis, personality traits, and COVID-19 related factors and opinions are presented in Supplementary Item S4.

**Item S4.**
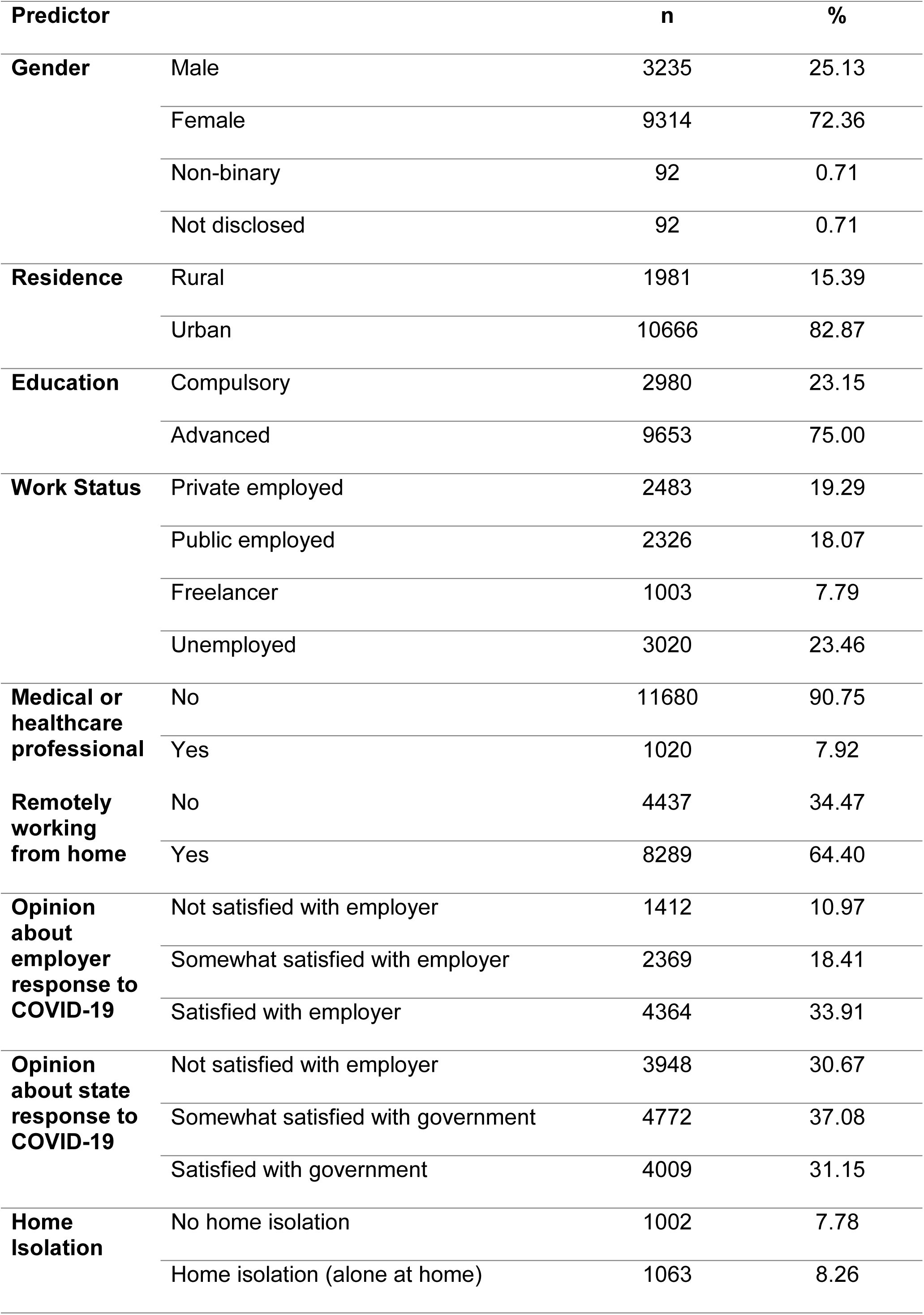
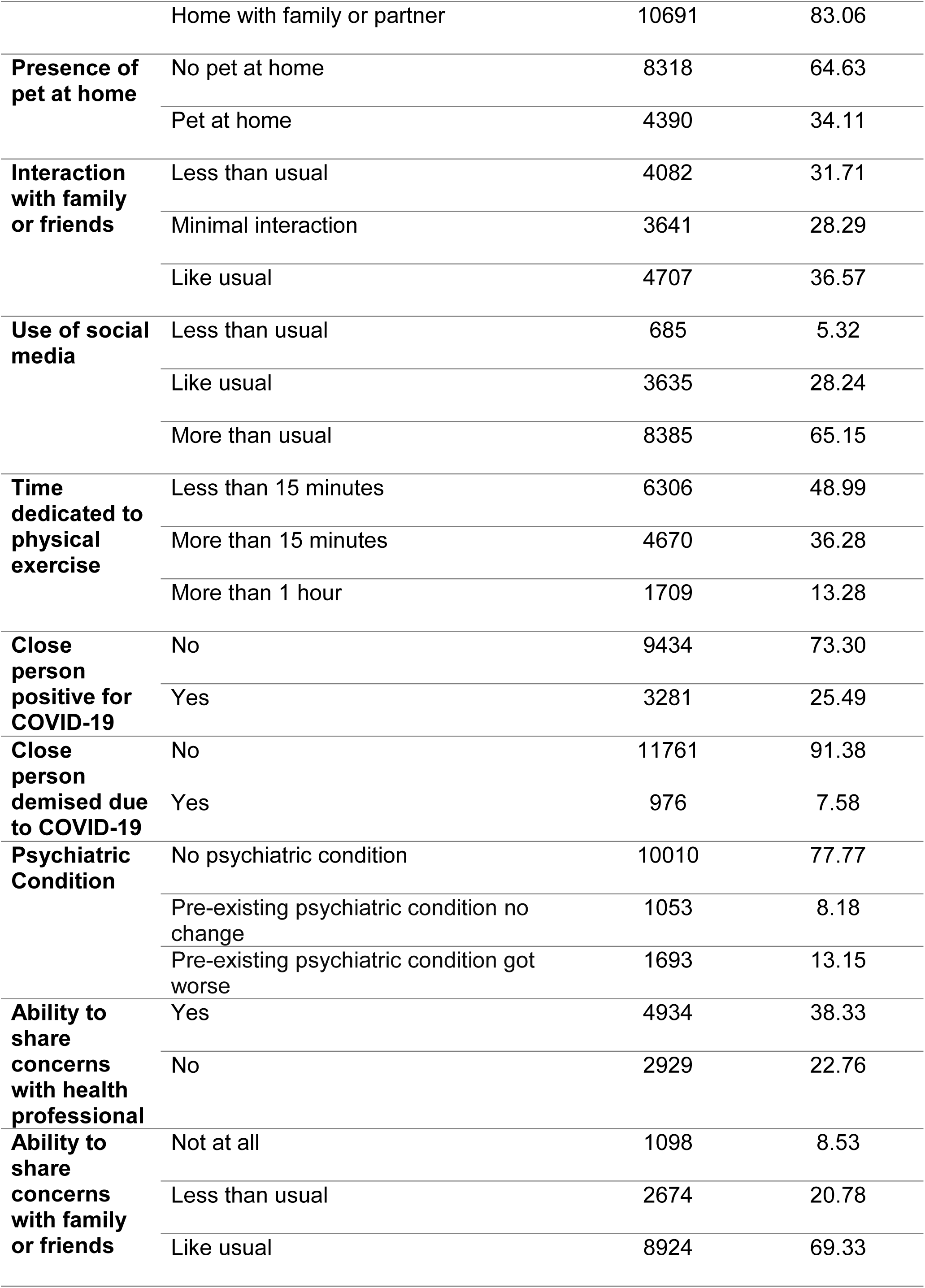
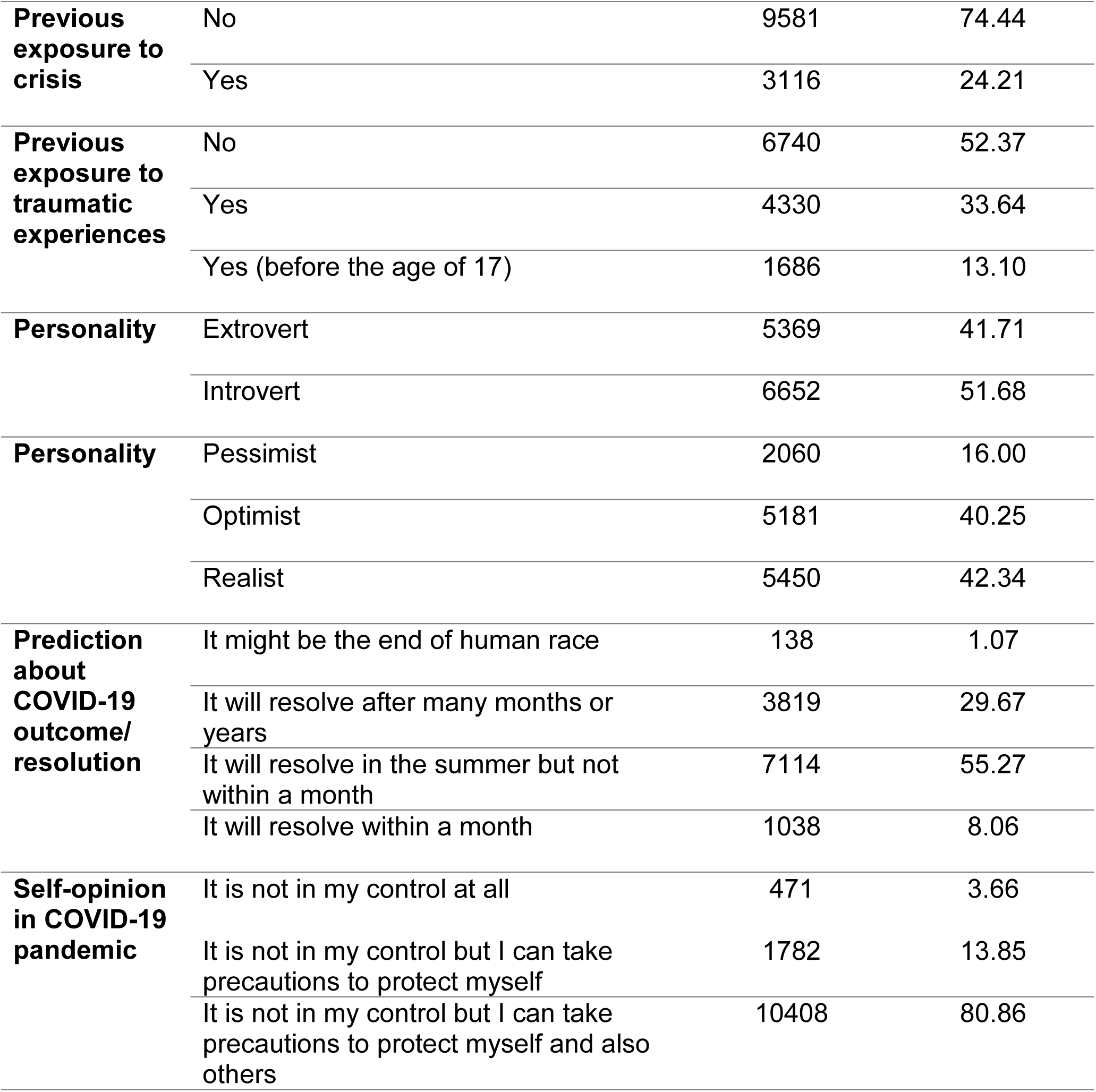
Table showing demographics and characteristics of the participants

### Unadjusted Analysis of Risk and Resilience Factors for General Psychological Disturbance (SRQ), PTSD Risk (IES), and Depression (BDI)

Unadjusted analyses of SRQ, IES, and BDI scores between different participant demographics/characteristics showed significantly (p<0.017) greater prevalence of psychological symptoms in participants who were female, unemployed, working remotely from home, dissatisfied with the response of their employer/state to COVID-19, home-isolated alone, with a pet, interacting with friends/famiy less than usual, using social media more than usual, and in those with less-than-usual ability to share concerns with friends/family. Significantly (p<0.017) higher scores on SRQ, IES, and BDI were also seen in participants who self-reported as being pessimist or introvert, not feeling in control during COVID-19, and having an overall negative prediction about COVID-19 resolution. Means and standard deviations for all comparisons are presented in Main Item 2.

**Main Item 1:**
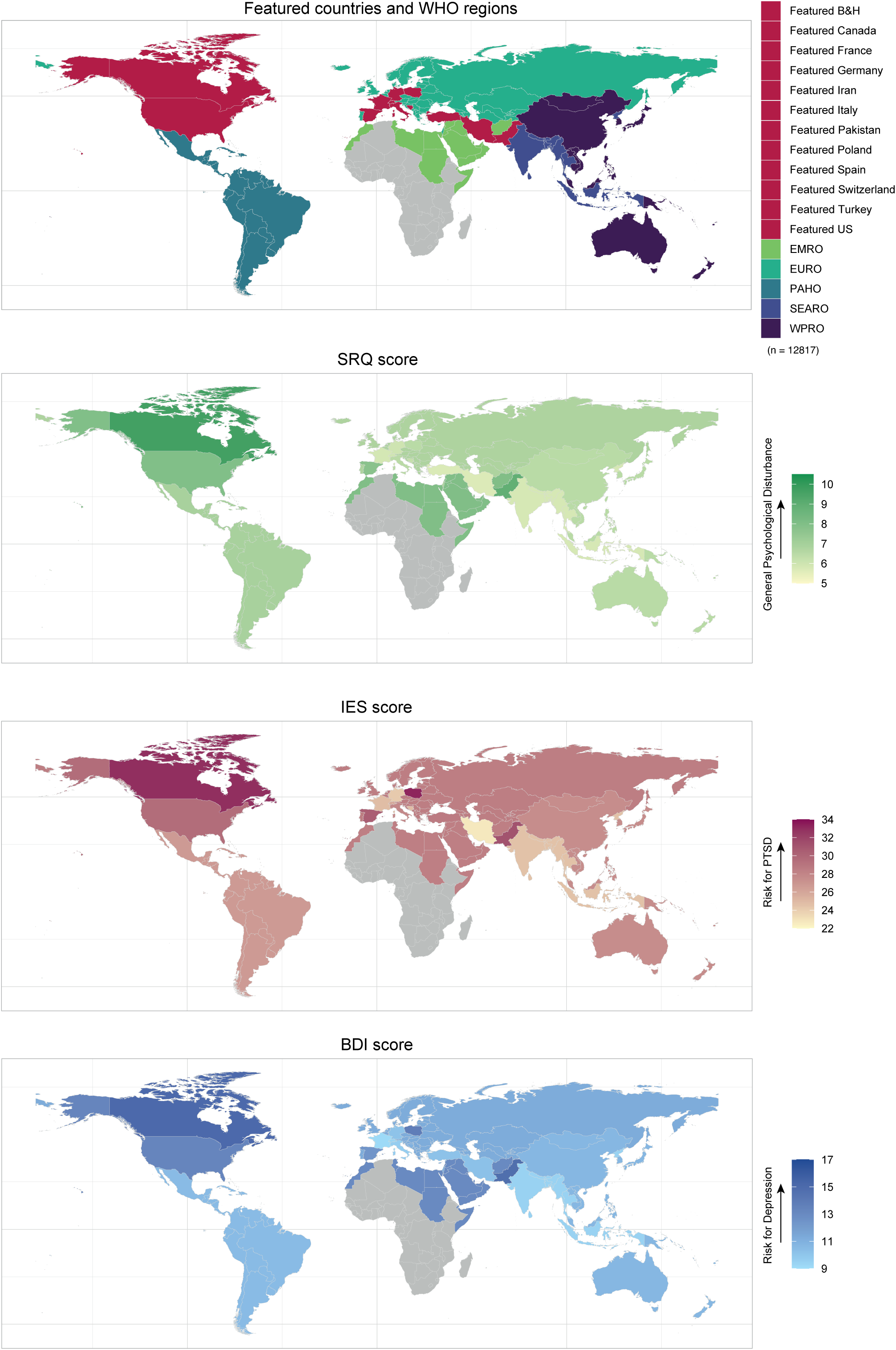
Geodemographic representation of global mental health burden. First map presents featured countries (red) and remaining countries divided according to WHO regions. Remaining maps present mean scores from SRQ, IES and BDI respectively. The means were calculated separately for each of the featured countries, and for each of the WHO regions. Total number of responders is 12,817, distributed as follows: USA (1864), Iran (1198), Pakistan (1173), Poland (1110), Italy (1096), Spain (972), Bosnia and Herzegovina (885), Turkey (539), Canada (538), Germany (534), Switzerland (489), France (337), and in remaining countries from WHO regions: WHO European region EURO (784), WHO East Mediterranean region EMRO (459), WHO Western Pacific region WPRO (326), WHO South East Asia region SEARO (259) and WHO region of the Americas PAHO (254). First panel: featured countries (in red) and remaining countries divided according to WHO regions; Second panel: mean scores for Self-Reporting Questionnaire (SRQ) indicating general psychological disturbance; Third panel: mean scores for Impact of Event Scale (IES) indicating risk for post-traumatic stress disorder (PTSD); Fourth panel: mean scores for Beck’s Depression Inventory (BDI) indicating risk for depression. All mean scores were calculated separately for the featured countries and WHO regions.

**Main Item 2:**
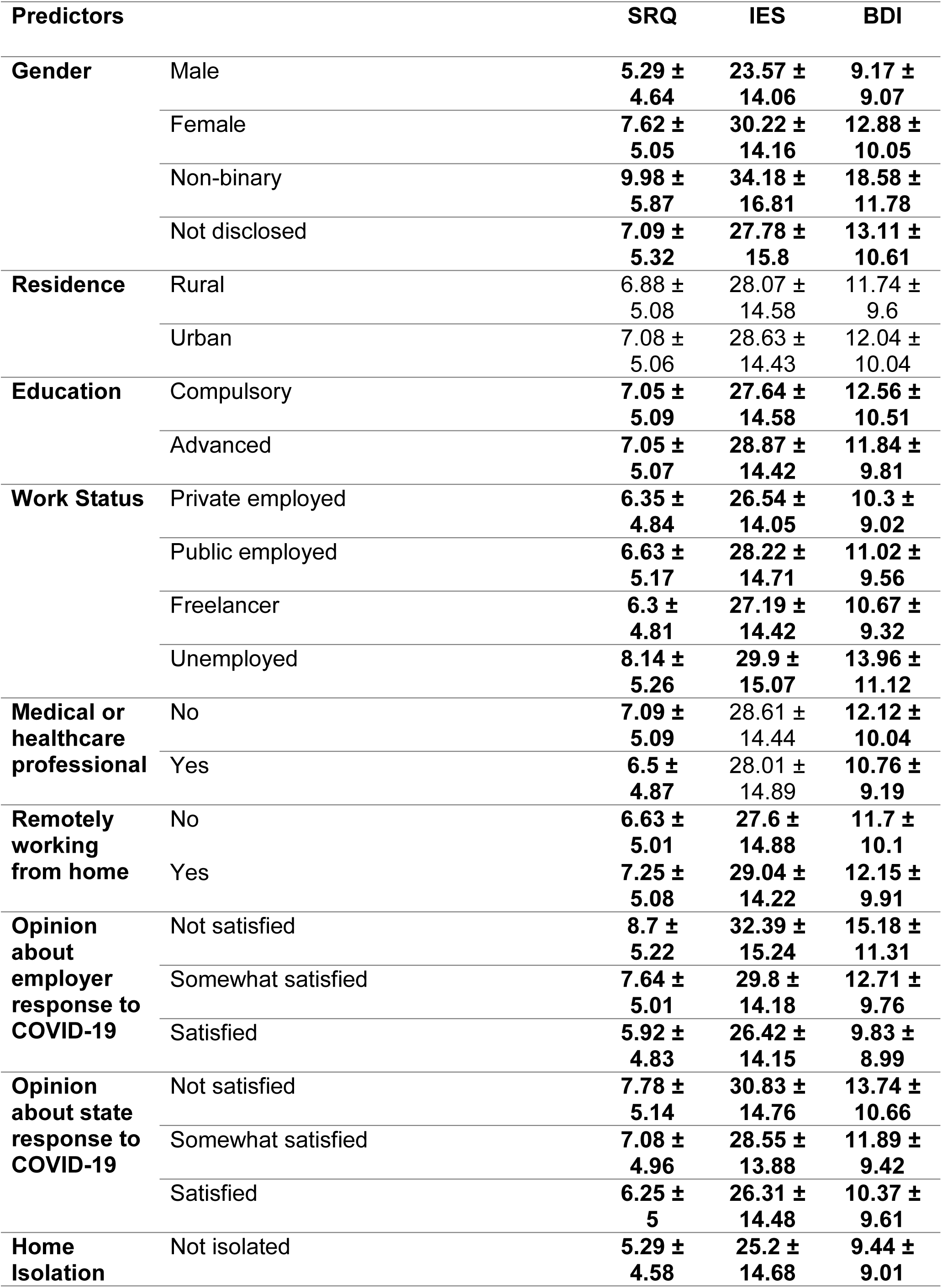
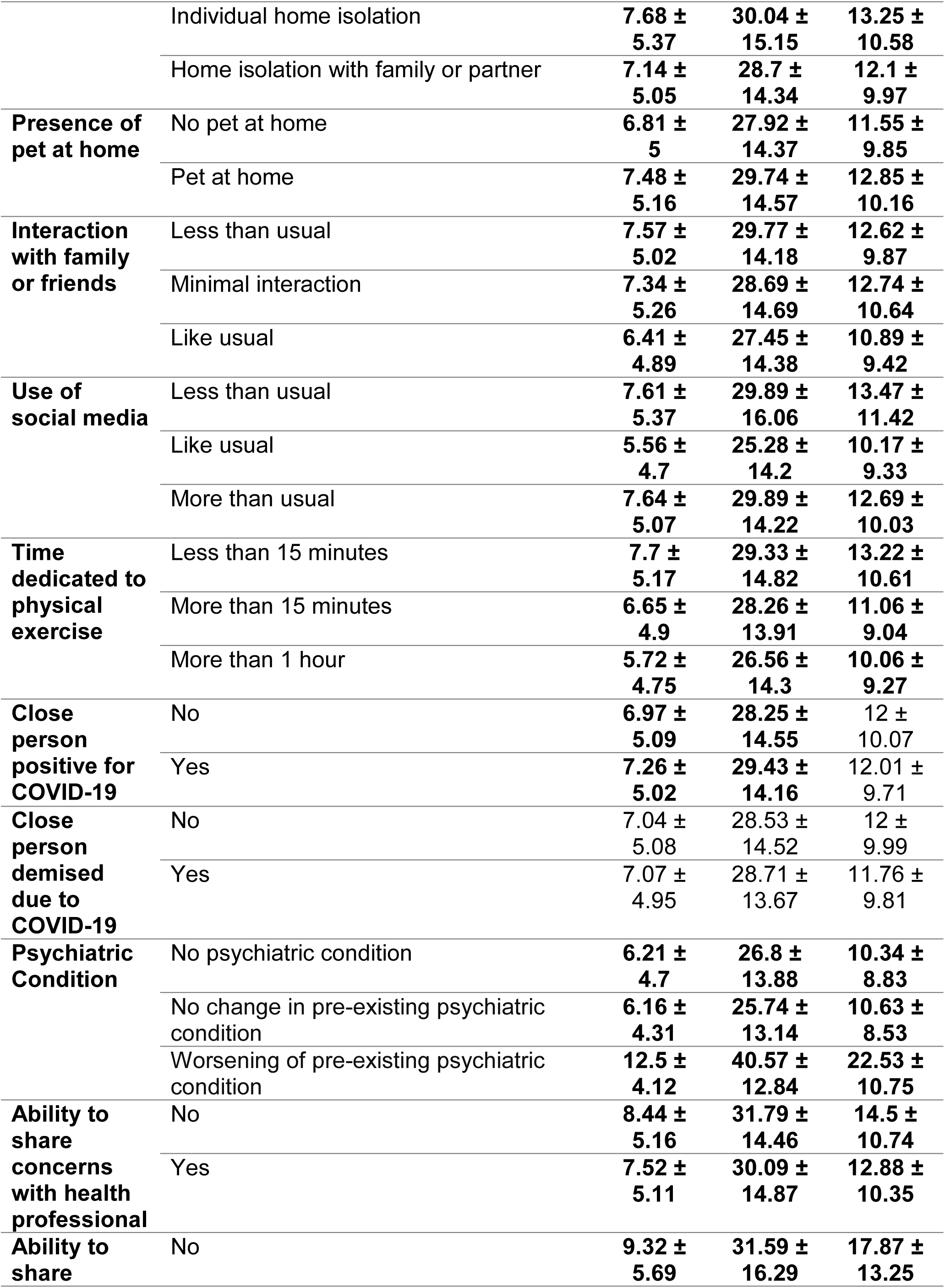
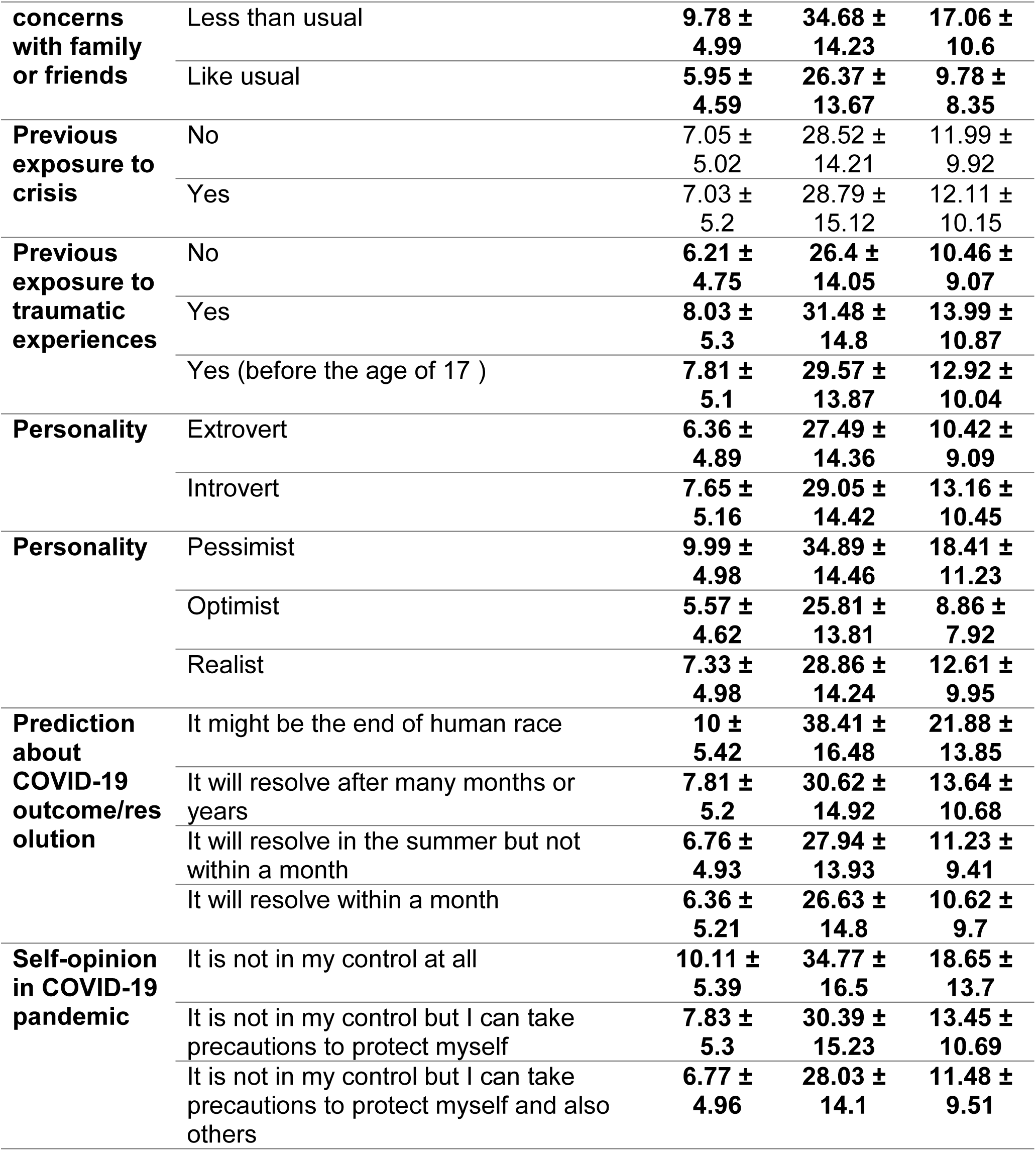
Comparison of psychological symptoms between different participant demographics/characteristics. This table shows means ± standard deviations of participants’ SRQ, IES, and BDI scores divided according to different participant demographics/ characteristics and compared through unadjusted Kruskal-Wallis tests. Significant differences (p-value threshold set to <0.017 after multiple-comparisons correction) in mean scores are highlighted as bold. Each bold association indicates difference in categories reported in the predictors column vertically.

### Adjusted Analysis of Risk and Resilience Factors for General Psychological Disturbance (SRQ), PTSD Risk (IES), and Depression (BDI)

Adjusted analysis using different general linear models for each of the questionnaires is reported in Main Item 3. Across all three questionnaires, we found the following relevant risk factors for general psychological disturbance, PTSD, and depression: psychiatric condition that worsened during the COVID-19 pandemic (SRQ mean-coefficient: 0.36, 95 % CI: [0.33, 0.39]; IES mean-coefficient: 7.36 95 % CI: [6.26, 8.46]; BDI mean-coefficient: 0.38, 95 % CI: [0.36, 0.40]), previous exposure to trauma (SRQ mean-coefficient: 0.19, 95 % CI: [0.16, 0.22]; IES mean-coefficient: 4.08 95 % CI: [3.14, 5.03]; BDI mean-coefficient: 0.20, 95 % CI: [0.17, 0.22]) and working remotely from home (SRQ mean-coefficient: 0.07, 95 % CI: [0.05, 0.10]; IES mean-coefficient: 1.91, 95 % CI: [1.01, 2.82]; BDI mean-coefficient: 0.03, 95 % CI: [0.01, 0.05]).

**Main Item 3:**
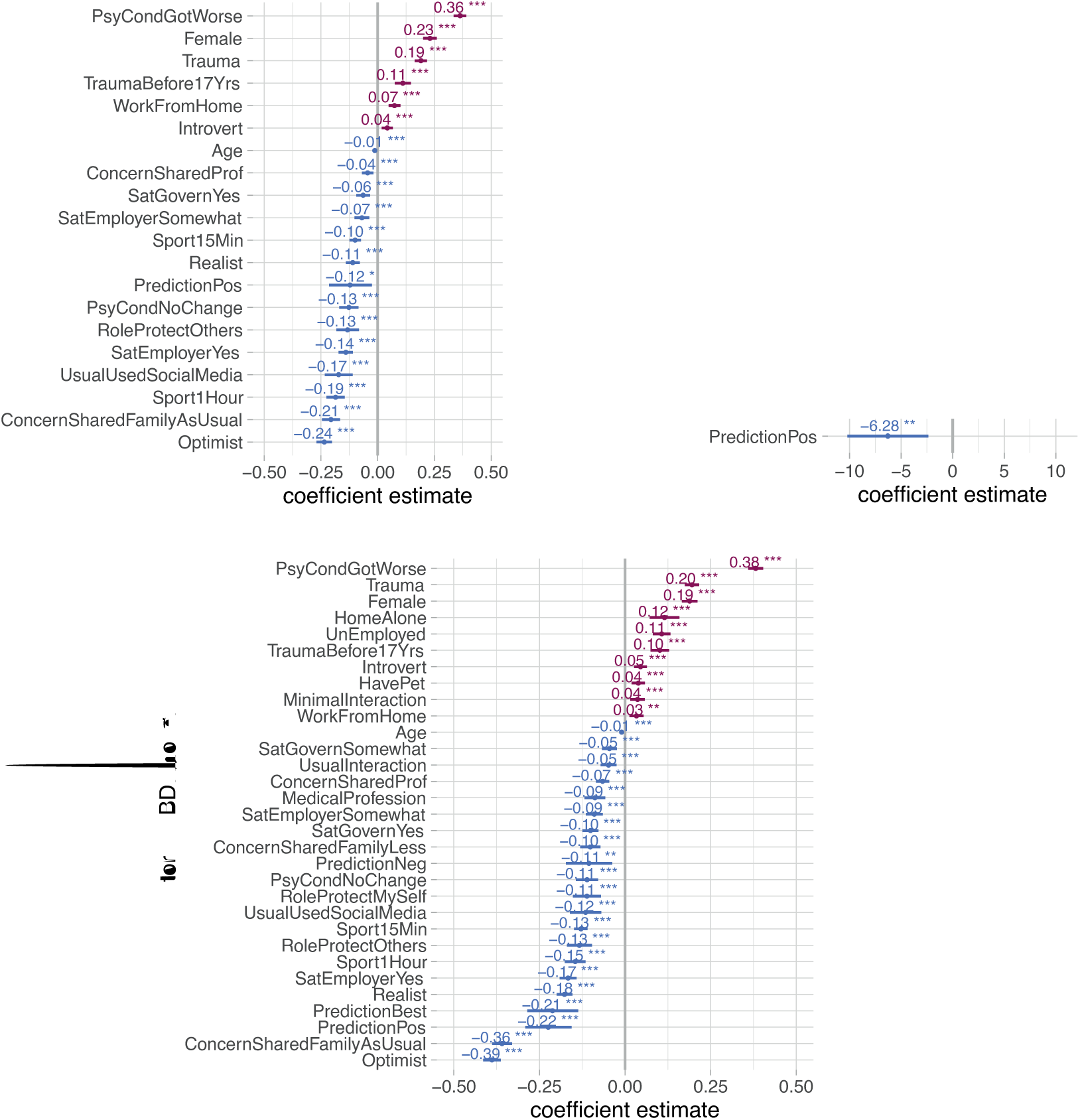
Risk and resilience factors for general psychological disturbance (SRQ), risk for PTSD (IES), and depression (BDI) These foster-plots show the mean estimates and the 95% confidence intervals (CI) for adjusted coefficients significantly affecting SRQ, IES and BDI scores respectively generated through multiple regression models. Only predictors that survived Bonferroni correction for multiple comparisons (p<0.017) are listed. Risk associations (i.e. increase in scores) are shown in red while resilience associations (i.e. decrease in scores) are in blue.

Moreover, significant gender differences were observed, with higher risk in women versus men for general psychological disturbances (SRQ mean-coefficient: 0.23, 95 % CI: [0.20, 0.26]), PTSD (IES mean-coefficient: 4.99, 95 % CI: [4.03, 5.95]), and depression (BDI mean-coefficient: 0.19, 95 % CI: [0.17, 0.21]).

Having an optimistic attitude, positive prediction about COVID-19, and being able to share concerns with family/friends decreased SRQ, IES, and BDI scores, indicating the protective effect of these factors for general psychological disturbance, PTSD and depression (as shown in Main Items 3 and 4). Furthermore, daily physical activity/sport decreased both SRQ (mean-coefficient: -0.19, 95 % CI: [-0.23, -0.15]) and BDI (mean-coefficient: -0.15, 95 % CI: [-0.18, -0.12]) scores, with greater protective effect with higher duration of the physical activity/sport (exercise ≥ 1 hour more effective in decreasing SRQ and BDI scores compared to exercise >15 minutes but <1 hour). In addition, healthcare professionals reported significantly lower BDI scores, suggesting this status to have a protective effect against depression (mean-coefficient: -0.09, 95 % CI: [-0.12, -0.06]).

**Main Item 4:**
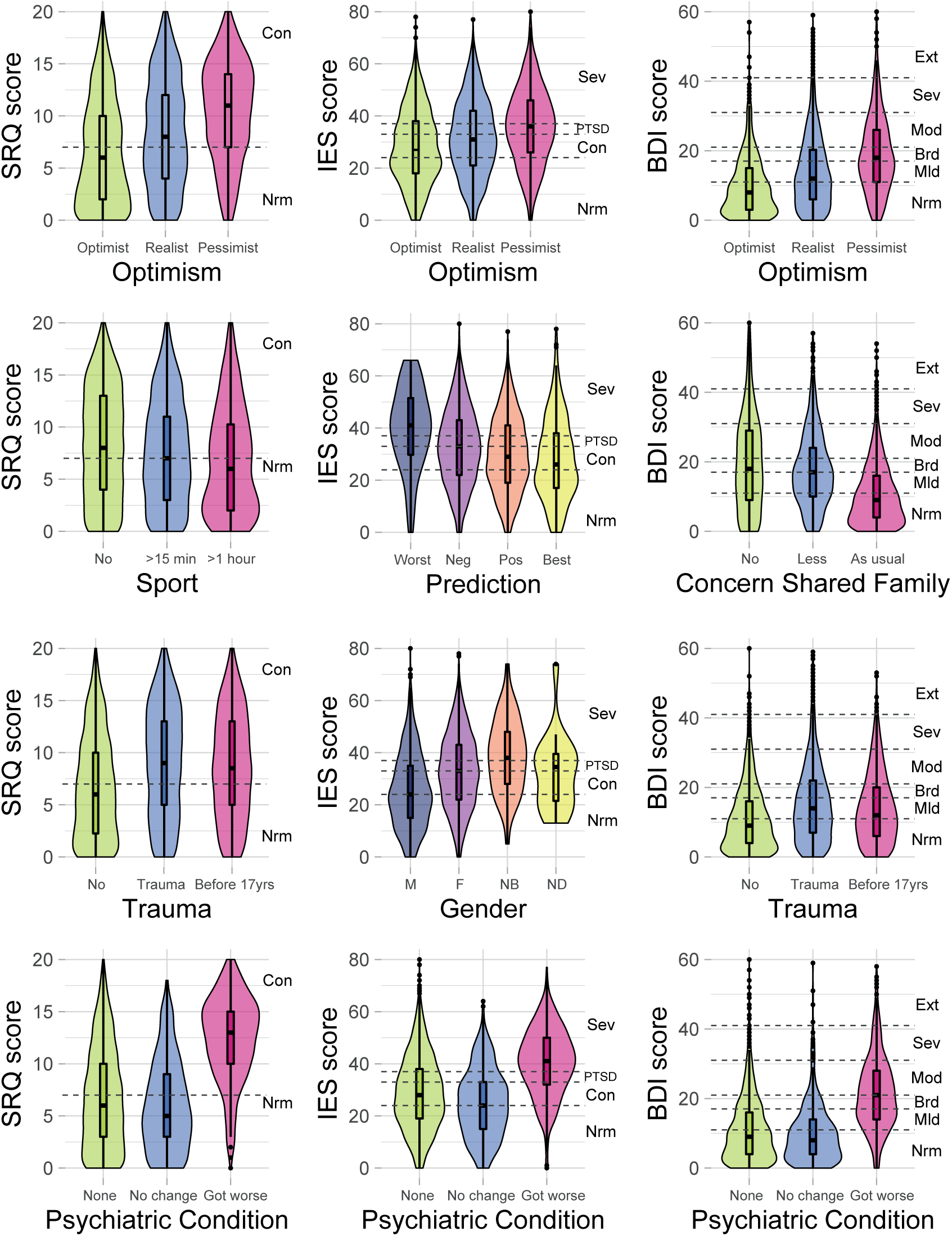
Violin plots indicating the effects of selected predictors on general psychological disturbance (SRQ), risk for PTSD (IES), and depression (BDI) These plots provide a relation between the participant scores on SRQ, IES, and BDI; and participant characteristics (previous history of a psychiatric condition, past exposure to trauma, prediction about COVID-19 resolution, and level of optimism, gender and daily physical activity/sport adjusted for confounding variables through multiple regression models. Boxplots display the distribution of the selected predictors with the visualization of five summary statistics (minimum, maximum, median, first quartile, third quartile), and all outliers individually. Violin plots added behind the boxplots visualize the probability density of selected predictors. Parallel to the x-axis, dashed lines present cut-offs for the scales used- For BDI: Ext - “Extreme”, 40 + points, Extreme Depression; Sev - “Severe”, 31-40 points, Severe Depression; Mod- “Moderate”, 21-30 points, Moderate Depression; Brd - “Borderline”, 17-20 points, Borderline clinical depression; Mld - “Mild”, 11-16 points, Mild mood disturbance; Nrm - “Normal”, 1-10 points, Considered normal For SRQ: Con - “Concern”, 11-20 points, Clinical concern for General Psychological Disturbance; Nrm - “Normal”, 0-10 points For IES: Sev- “Severe”, 37+ points, symptoms high enough to suppress the immune system; PTSD - “Post Traumatic Stress Disorder”, 34-36 points; Con - “Clinical Concern for possible PTSD”, 24-33 points, Nrm - “Normal”, 0-23 points

The logistic regression analyses performed after classifying SRQ, IES, and BDI scores into categorical cut-offs confirmed the primary results from the linear regression models (Supplementary Item S5). An individual with pre-existing psychiatric condition that worsened during COVID-19 showed 7-times higher odds of being depressed (OR: 7.10, 95% CI: [6.03, 8.35]), 1.6 times higher odds of having PTSD (OR:1.60, 95% CI: [1.38,1.84]) and twice higher odds of having general psychological disturbance (OR: 2.64, 95% CI: [1.99,3.48]). As expected, individuals with previous trauma exposure exhibited greater ORs than their counterpart for these conditions according to BDI (OR: 1.61, 95% CI: [1.46, 1.76]) and SRQ (OR: 2.62, 95% CI: [2.08, 3.30]). Still, an optimistic attitude and the opportunity to share concerns with family/friends like usual served as a protective factor for general psychological disturbance according to SRQ (OR: 0.51, 95% CI: [0.43, 0.62] and OR: 0.19, 95% CI: [0.15, 0.23] and depression according to BDI (OR: 0.23, 95% CI: [0.20, 0.26] and OR: 0.39, 95% CI: [0.33, 0.45] respectively.

**Item S5.**
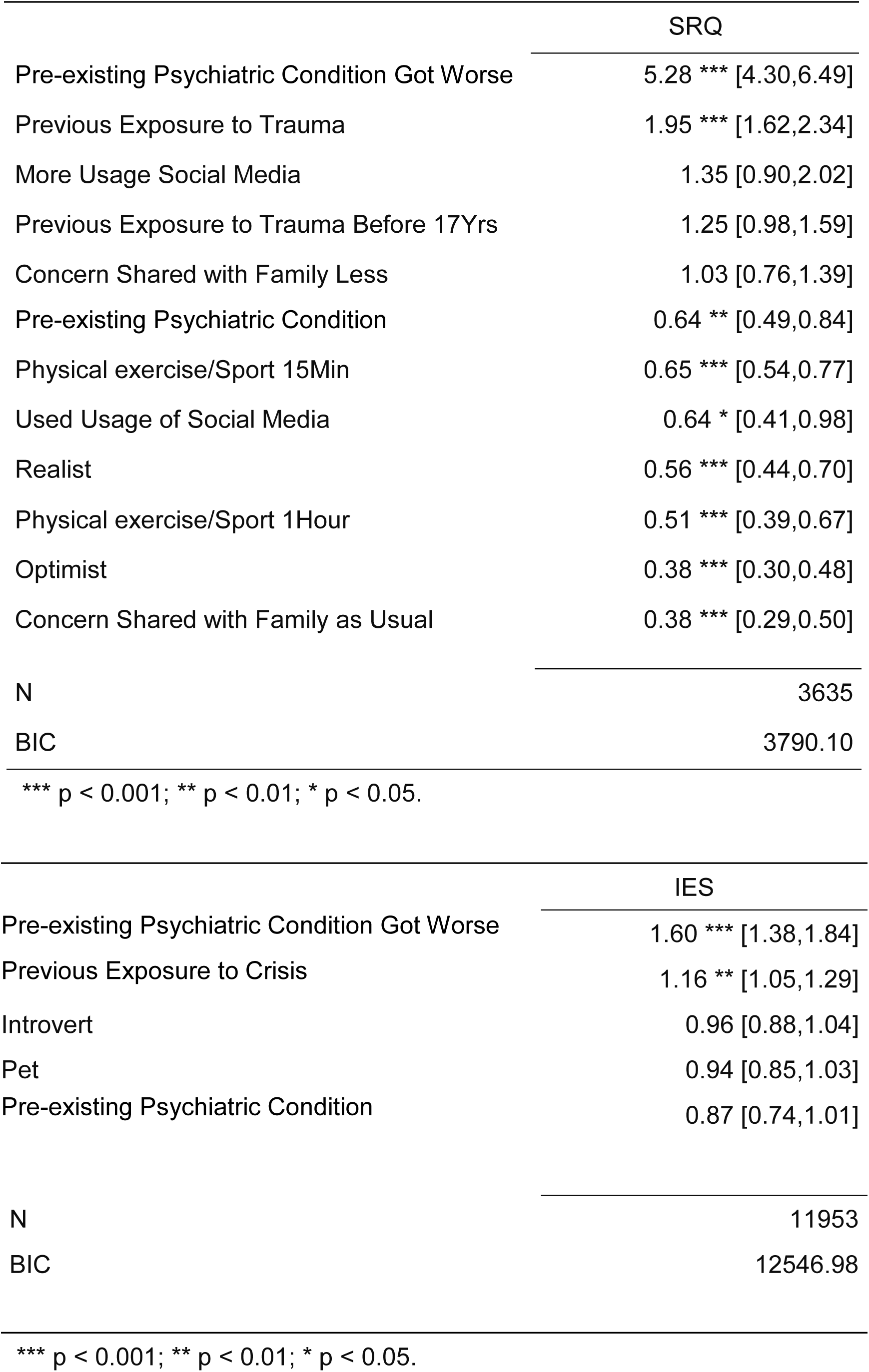
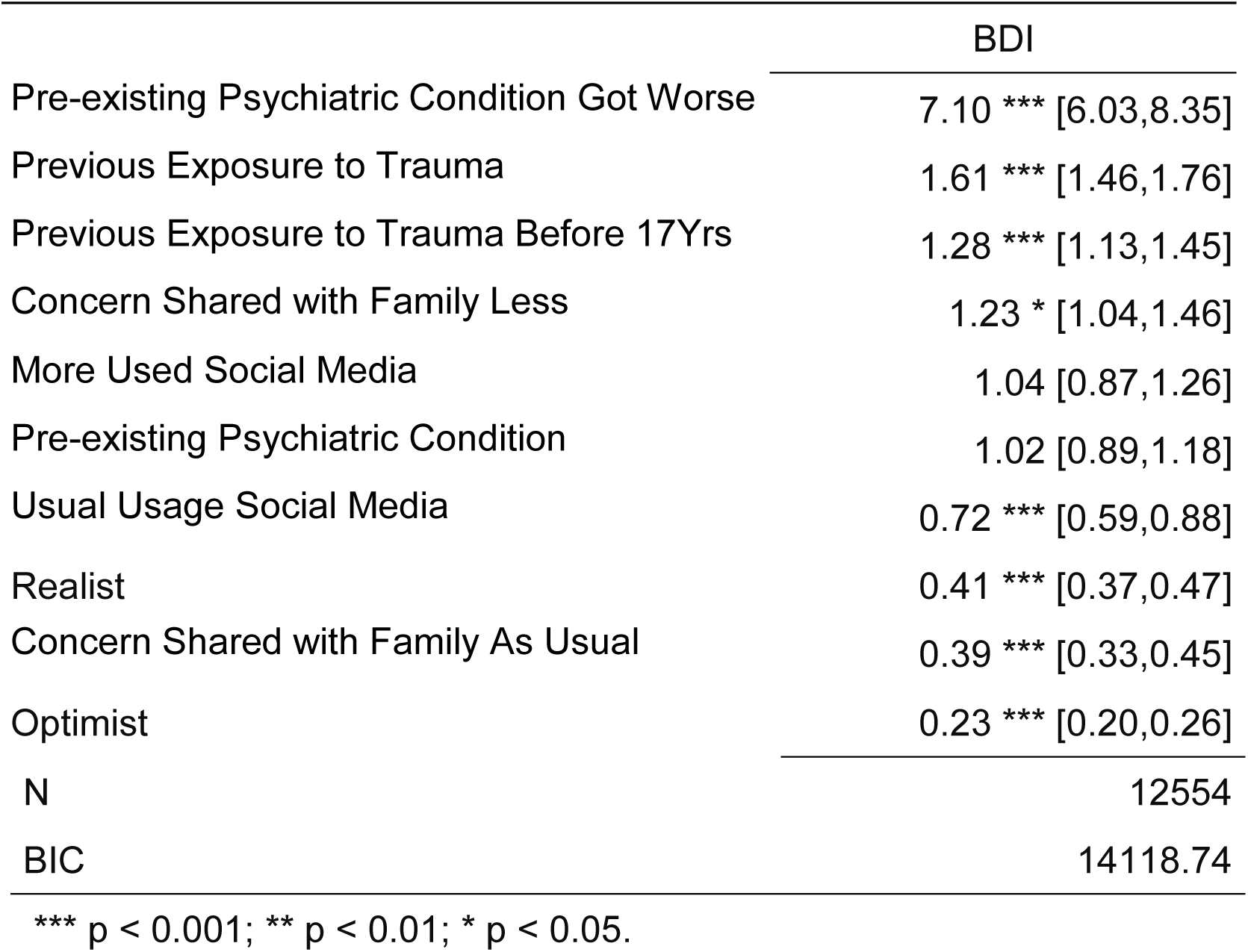
Predictors for general psychological disturbance, PTSD, and depression. Logistic regression was performed to generate odds ratios (ORs) for SRQ, IES, and BDI using the following categorization scheme; SRQ: 0 = normal (0-7 points), 1 = concern for general psychological disturbance (8-20 points); IES: 0 = normal (0-23 points), 1 = PTSD is a clinical concern (24-32 points), 2 = threshold for a probable PTSD diagnosis (33-36 points), 3 = Severe condition (high enough to induce immunosuppression) (37+ points). For generating ORs, the variables were regrouped as 0 = no concern versus any type of concern (1/2/3); BDI: 0 = These ups and downs are considered normal (1-10 points). 1 = Mild mood disturbance (11-16 points), 2 = Borderline clinical depression. (17-20 points), 3 = Moderate Depression (21-30 points), 4 = Severe Depression (31-40 points), 5 = Extreme Depression (>40 points). For generating ORs, the variables were regrouped as 0 = no concern versus any type of concern (levels 1/2/3/4/5). Only the factors that survived step BIC models comparison are listed. OR >1 indicate increased risk and OR<1 indicates protective effect.

For the ease of understanding, the association of participant-related predictors with categorical classifications for general psychological disturbance (SRQ), PTSD (IES), and depression (BDI) are indicated through box-plots in Main Item 5. Owning a pet, pre-existing psychiatric condition, previous exposure to trauma, considering oneself an introvert, and working remotely from home were associated with decreased %age of responses in the unaffected (‘normal’) category based on SRQ, IES, as well as BDI, suggesting these as risk factors. Contrastingly, a majority of responses from health professionals landed in the unaffected (‘normal’) category for BDI, indicating that working as a health professional is a resilience factor against depression during the COVID-19 pandemic.

**Main Item 5.**
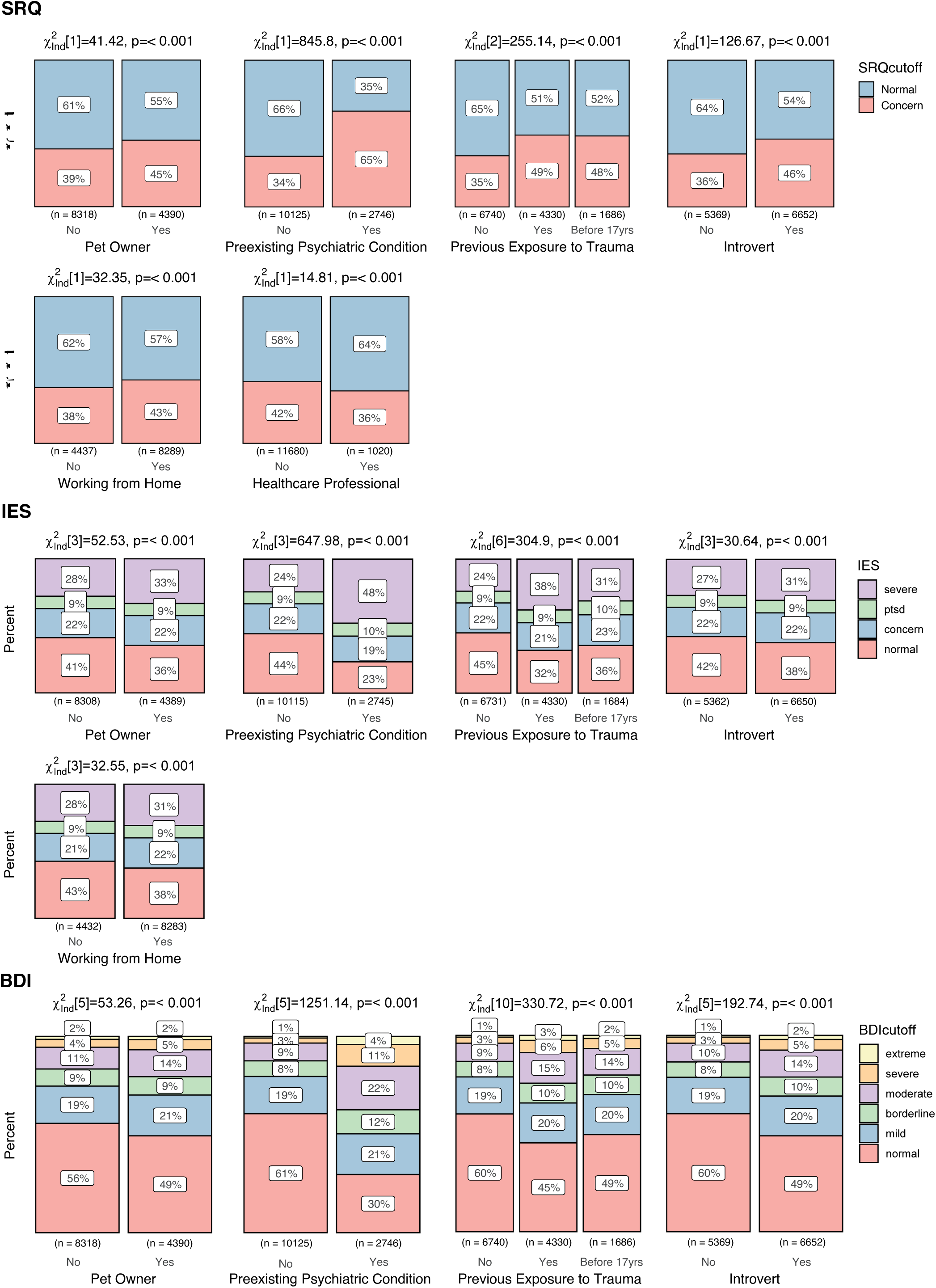
Association of participant demographics/characteristics and categorical classifications for general psychological disturbance (SRQ), PTSD (IES) and depression (BDI) These tables show results from the predictors with a significant chi-square value after correcting for multiple comparisons (Bonferroni correction for 27 tests, p<0.0019). For each predictor, the bar plot indicates the participant’s distribution across the different categories of SRQ, IES, and BDI. The cut-offs used are as follows: for SRQ normal/concern (0-17, 8-20 points); for IES normal/concern/PTSD/severe (0-23, 24-32, 33-36, 37+ points); for BDI normal/mild/borderline/moderate/severe/extreme (1-10, 11-16, 17-20, 21-30, 31-40, 40+ points). The plots include the total number and %age of participants in each category and the statistical outcomes from the chi-square test.

### Suicidal Ideation and Concerns for Physical Health and Appearance

Responses to three relevant questions from BDI indicating suicidal ideation and concerns about physical health and physical appearance were analyzed separately through logistic regression models.

Worsening of pre-existing psychiatric condition and past exposure to trauma predicted increased suicidal ideation (OR: 4.66, 95% CI: [4.10, 5.29] and OR: 1.56, 95% CI: [1.38, 1.75]; Supplementary Item S6). On the contrary, ability to share concerns with family and friends like usual and optimistic attitude decreased suicidal ideation by almost 70% percent (OR: 0.30, 95% CI: [0.26,0.36] and OR: 0.32, 95% CI: [0.27,0.36] respectively). Similarly, a pre-existing psychiatric condition that worsened during COVID-19 increased the likelihood of having concerns about both physical health and physical appearance (OR: 2.80, 95% CI: [2.49, 3.14] and OR:2.85, 95% CI: [2.52, 3.22] respectively; Supplementary Items S7 and S8). In addition, individuals with previous trauma exposure were more likely to show increasing concerns about their physical health (OR: 1.56, 95% CI: [1.43, 1.69]). Moreover, the odds for women being concerned about physical appearance were about 70% higher as compared to men (OR: 1.70, 95% CI: [1.55, 1.87], Supplementary Item S8).

**Item S6.**
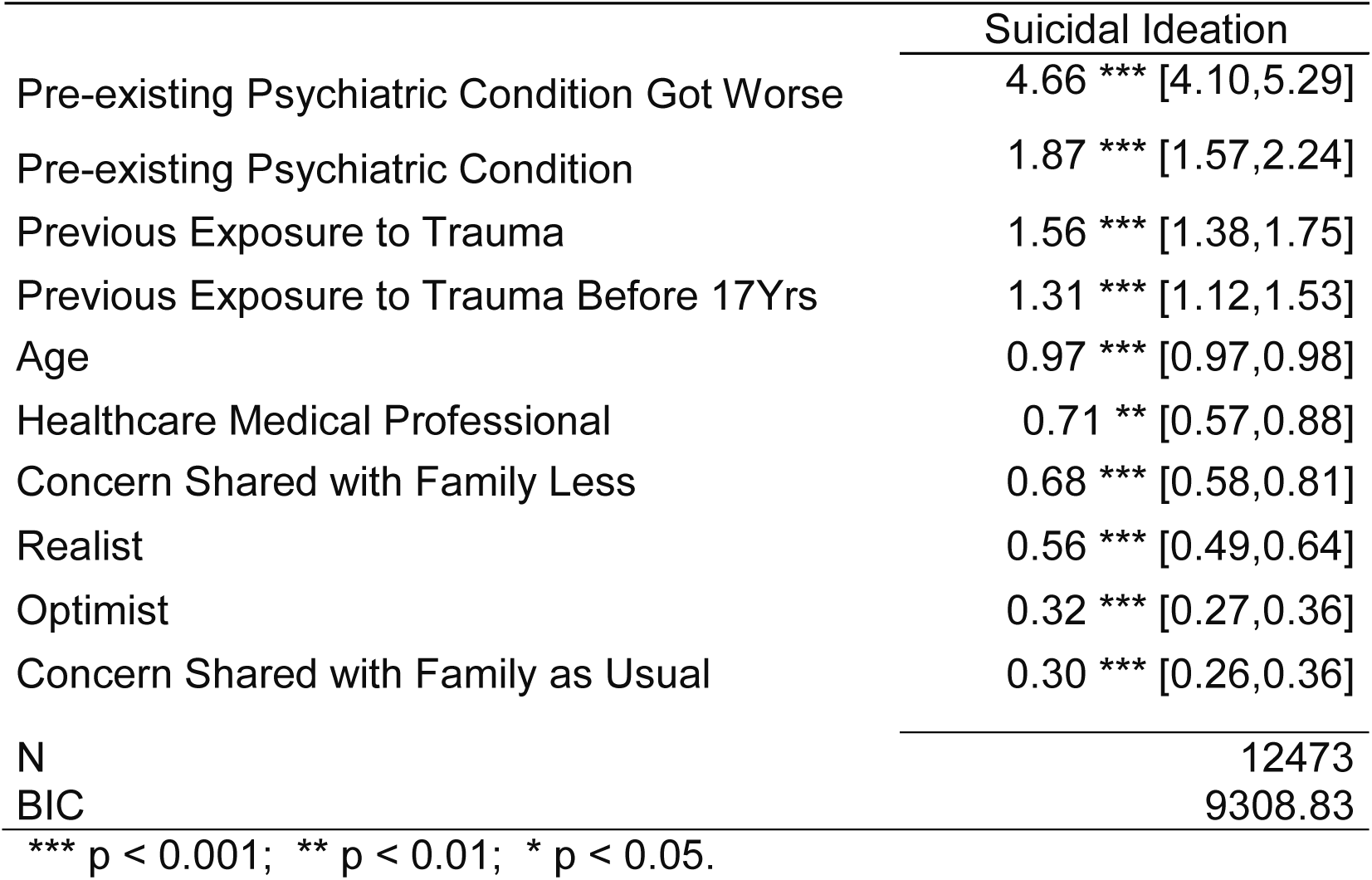
Predictors for suicidal ideation. Presentation of the odds ratios (ORs) for suicidal ideation based on the relevant question from BDI. BDI question screening suicidal ideation is rated on likert-type scale (0-3) indicating increasing suicidal ideation. For this analysis, logistic regression was performed regrouping the variables as 0 = no suicidal ideation versus any suicidal ideation (1/2/3). Only the factors that survived step BIC models comparison are listed. OR >1 indicates risk and OR<1 indicates protective effect.

However, daily physical exercise/sport for one hour or more, as well as having an optimistic attitude decreased the odds for concerns about physical appearance by around 30% and 60% respectively (OR: 0.61, 95% CI: [0.54, 0.68] and OR: 0.37, 95% CI: [0.33, 0.42]], Supplementary Item S8). Those two factors were also protective against concerns about physical health by about 50% (OR: 0.57, 95% CI: [0.50, 0.64] and OR: 0.56, 95% CI: [0.50, 0.62], Supplementary Item S7). We note the essential role of physical exercise, demonstrated in multiple areas of this study, in promoting a positive outlook on one’s health and appearance.

**Item S7.**
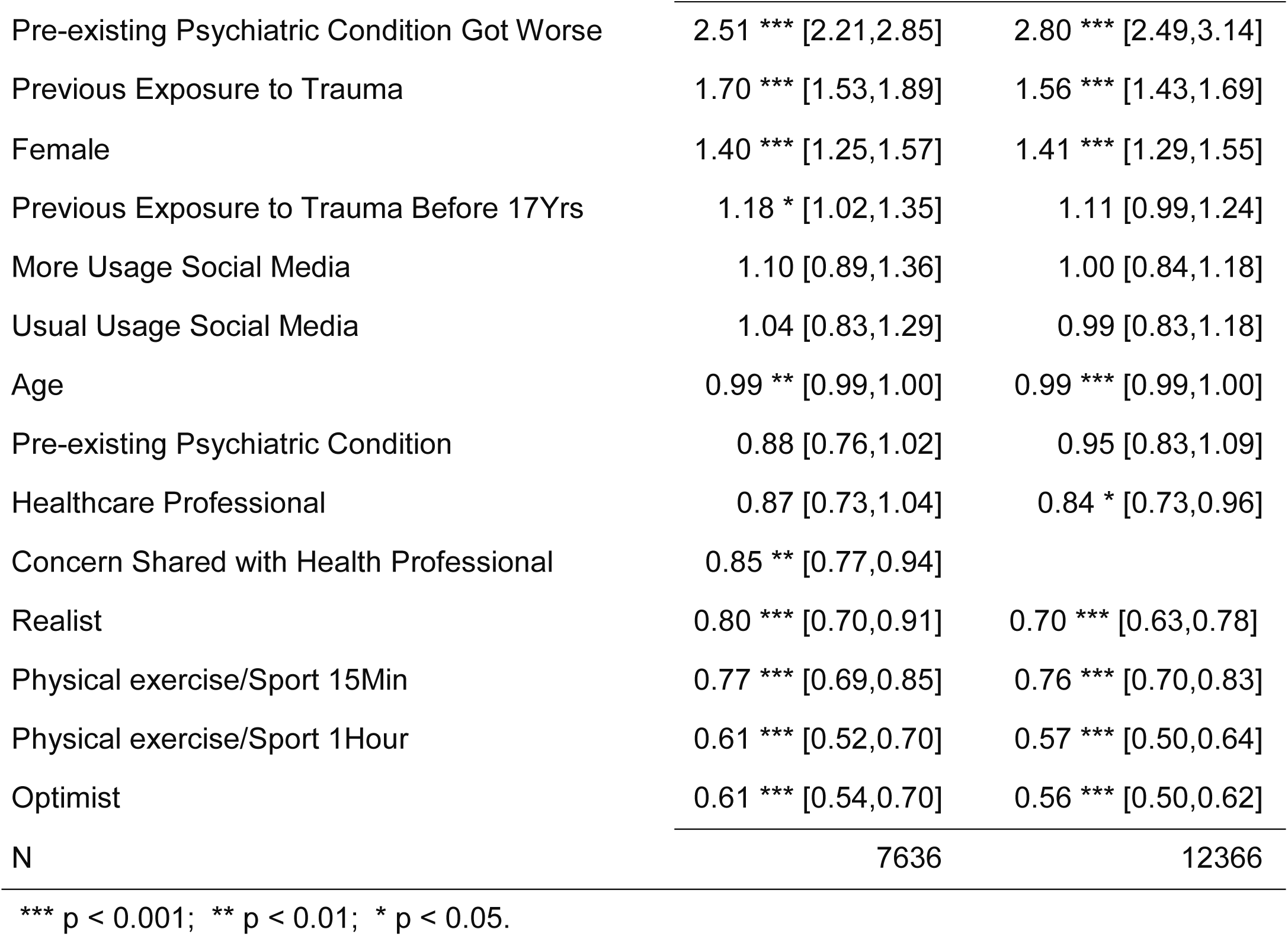
Predictors for concern about physical health. Presentation of odds ratios for concern about physical health based on the relevant question from BDI. BDI question screening concern for physical health is rated on likert-type scale (0-3) indicating increasing concern for physical appearance. For this analysis, logistic regression was performed regrouping the variables as 0 = no concern versus any concern (1/2/3). Only the factors that survived step BIC models comparison are listed. OR >1 indicates risk and OR<1 indicates protective effect.

**Item S8.**
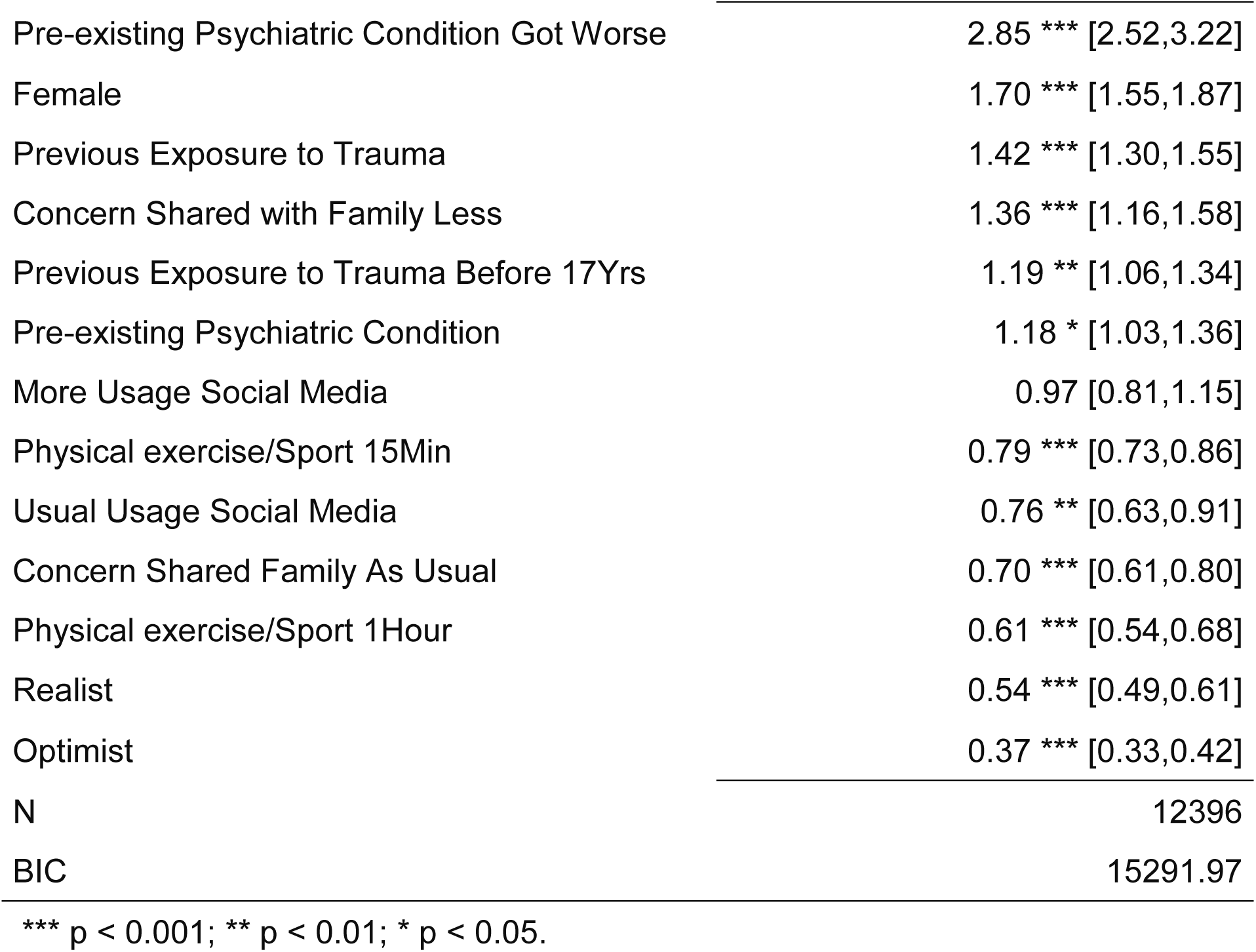
Predictors for concern about physical appearance. Presentation of odds ratios for concern about physical appearance based on the relevant question from BDI. BDI question screening concern for physical appearance is rated on likert-type scale (0-3) indicating increasing concern for physical appearance. For this analysis, logistic regression was performed regrouping the variables as 0 = no concern versus any concern (1/2/3). Only the factors that survived step BIC models comparison are listed. OR >1 indicates risk and OR<1 indicates protective effect.

### Correlation between Scales

The continuous scores of all responses on SRQ, BDI, and IES were also analyzed by Pearson’s correlations using all possible combinations on x~y plotting (SRQ vs. IES, IES vs. BDI, BDI vs. SRQ). All combinations yielded significant correlations with the strongest correlation (R=0.79) between BDI and SRQ (Supplementary Item S9).

**Item S9.**
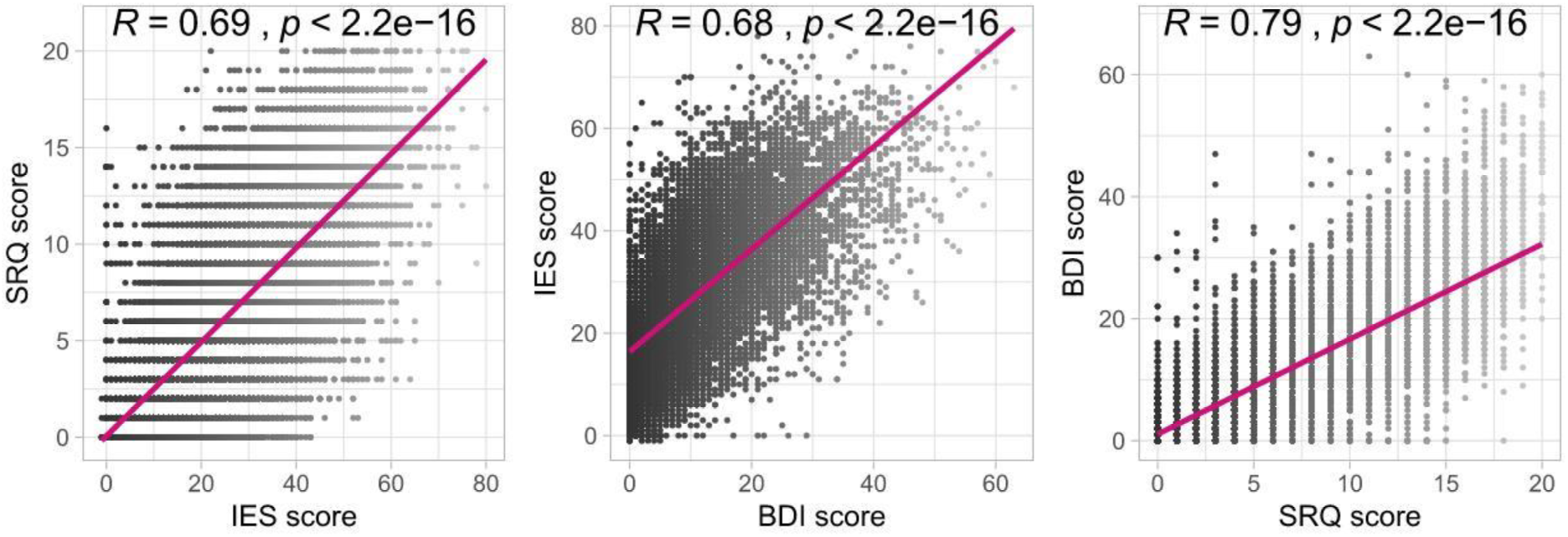
Correlations between the SRQ, IES, and BDI scores. These plots show Pearson’s correlation values for all the possible pairs of the three scales used, i.e., SRQ vs. IES, IES vs. BDI, and BDI vs. SRQ. All the correlations are statistically significant and range from 68% to 79% correlation.

## DISCUSSION

This study highlights a significant impact of COVID-19 pandemic on mental health worldwide. Baring a few outliers, participants across the 12 featured countries and other countries clustered into five WHO regions had scores exceeding the mild-risk threshold for general psychological disturbance, PTSD, and depression, as determined by standardized scales. Furthermore, an alarming fraction (16.2%) of the participants reported experiencing some level of suicidal ideation. A prominent fraction (41%) of the participants also expressed concerns about their physical health and appearance, which is known to accompany other forms of psychological distress.^34,38^

In addition to reporting prevalence, a major aim of this study was to identify specific risk and resilience factors for psychological perturbations during the current COVID-19 crisis. Worsening of a pre-existing psychiatric condition, female gender, exposure to trauma before age 17, and working remotely predicted higher risk of general psychological disturbance, PTSD, depression, and increased concerns about physical health and appearance. Additionally, considering oneself an introvert was associated with heightened risk of general psychological disturbance and depression; being unemployed, living alone, and limited interaction with family and friends also increased the risk for depression. Pre-existing psychiatric conditions and previous exposure to traumatic events predicted suicidal ideation. An overall protective effect against all major psychological perturbations was observed for the following factors; increasing age, considering oneself an optimist, positive prediction about COVID-19 outcome, ability to share concerns with family and friends like usual, daily physical exercise/sport for 15 minutes or more, and being satisfied with the actions of employer/state in response to COVID-19. Furthermore, being a health professional was associated with lower general psychological disturbance, depression, suicidal ideation and concern for physical health, and like-usual social media usage was associated with less concern for physical appearance.

To the best of our knowledge, this study is the first worldwide assessment of the mental health effects of COVID-19. Previous studies on the psychological impact of COVID-19 have been exclusively from China^13,19-25^ with the exception of one study in India.^39^ The largest of these studies (n=52,730) that surveyed voluntary public participants, reported symptoms of psychological distress in almost one-third of the participants according to the peri-traumatic distress index.^40^ Another notable study, on health professionals (n=1,255), revealed depression, anxiety, and symptoms of general distress in almost half of participants, and sleep disturbances in almost 8%.^13^ One-third of the participants in a Chinese study on college students (n=7,143) in the Hubei province reported symptoms of anxiety.^25^ Some of our observations are supportive of findings in these studies, such as female gender, living alone, and negative prediction about COVID-19 outcome arising as risk factors for psychological perturbations. However, our study identifies several unique risk and resilience factors that were not investigated previously.

Parallels can also be drawn between our study and existing research on the psychological effects of the SARS and other previous epidemics. These studies reported PTSD, anxiety, distress, anger, and confusion as major sequelae of the epidemic and quarantine measures.^35,41-43^ It has previously been reported, however, that very few studies investigated specific risk or protective factors^7^ for these mental health disturbances. One notable study showed longer quarantine duration, boredom, financial instability, stigma, inadequate resources and information deficit to exacerbate the negative psychological impact from the SARS outbreak. In noteworthy contrast to our work, the study was performed several months after the epidemic had occurred.^44^

Identification of specific risk and resilience factors is an essential first step for developing strategies to mitigate the negative psychological impact of COVID-19 at a regional and global level. For example, selective vulnerability of females indicated in this study warrants further investigation for both the contributing factors and the resulting implications of such increased risk. These include social factors such as increased reporting of domestic violence in relation to COVID-19 ^45^, possible caregiver stress, and the impact of changes in roles and responsibilities secondary to the current health emergency. Furthermore, increased risk of psychological perturbations in individuals with pre-existing psychiatric conditions and/or trauma exposure necessitates the initiation and/or expansion of mental health support systems available remotely.^40^ Emerging evidence now supports the efficacy of web- and social-media based interventions in promoting mental health of masses focusing on paradigms based on mindfulness, positive psychology, and exercise.^46-48^ Such interventions could be developed at the governmental and institutional levels and delivered to the masses via main-stream and social media. Indeed, media outlets could also play a major role in promoting optimism and a positive attitude towards COVID-19 resolution, both of which were identified in our study as important resilience factors. One example of media positivity could be reporting on ‘*number of active cases*’ rather than ‘*number of new cases*’, as this would also portray ‘*number of recoveries*’. It is of notable possibility that this method could not only serve to reduce psychological distress and hopelessness, but could also provide a more accurate estimate of the burden on health systems. Furthermore, the association between remote working with increased psychological symptoms calls for optimization of the work-from-home settings and a greater emphasis on the general well-being of employees. This is further corroborated by the observation that participant satisfaction with the employer-response to the COVID-19 pandemic is associated with reduced psychological symptoms in this study. Finally, the association of suicidal ideation with both pre-existing psychiatric condition and previous trauma exposure merits awareness efforts to inform the public about these risks. Such a finding could also warrant targeted interventions by mental health entities to mitigate the risk of suicide in these vulnerable populations.

An intriguing finding of this study is a mild protective effect of increasing age against general psychological disturbance, PTSD and depression. We recognize the possibility that this could be related to the study procedures. As the elderly are less comfortable with the use of electronic tools, a special provision was allowed for them to use assistance for recording their answers. However, this could have a confounding effect as older adults can be reluctant to openly report psychological symptoms.^49^ Additionally, the utility of one of our assessment tools, SRQ, in detecting psychological disturbances in the elderly has been previously challenged.^50^ Notably, a Chinese study also showed that adults aged 18-30 are most vulnerable to negative psychological effects of COVID-19^40^ as seen in a vast majority of our participants as well. Therefore, it is plausible that older adults are indeed less affected psychologically by COVID-19 because of their lesser reliance on social media, which another Chinese study found to be associated with increased anxiety and depression during the COVID-19 outbreak.^13^

This research has several strengths. This study employed the 2^nd^ largest sample size to date in examining the mental health impact of COVID-19, and the number of participants well exceeds previous studies on the SARS epidemic. The only study with a larger sample size^40^ employed a single scale for screening psychological disturbances. The administered measures in our study allowed for simultaneous screening of multiple psychiatric co-morbidities and the findings can provide invaluable insight to global health systems. The availability of the questionnaire in 11 different languages is a notable and unprecedented effort to provide the study as much generalizability as possible. Similarly, the 12 countries included in the featured list are not only representative of the different brackets of most severely affected countries globally, but also represent different economic strata. Canada, France, Germany, Italy, Spain, Switzerland, and USA are classified as high-income economies according to the World Bank Atlas, whereas, Bosnia and Herzegovina, Iran, Pakistan, and Turkey are middle or lower- income countries.^51^ Furthermore, the timing of this study is an important strength as the data was collected from March 29^th^-April 14^th^, 2020. This timing coincides with the peak of COVID-19 pandemic in North America and Europe—a time period when almost one-half of the world remained in complete lock-down.^4^ Finally, while cultural and linguistic factors are known to possibly impact psychological outcome measures when translations are utilized, the significant correlation between SRQ, IES, and BDI scores in this study cross-validates the assessment of psychological symptoms and confirms that COVID-19 pandemic is globally affecting the overall mental health of individuals.

The study also has potential limitations that warrant consideration when interpreting the results. First, the study employed a non-randomized sampling strategy. While this method has certain disadvantages, we hope that our results will catalyze the development of more studies on this essential topic that could be conducted by global outlets such as WHO and the European Union (EU) on a world- or continent-wide scale. Second, the data collection was exclusively done in an online format that may exclude those less-versed in web-usage, such as illiterate, disadvantaged, underdeveloped, or rural populations. We tried to reduce this bias by translating the study questionnaire into native/official languages for each of the featured countries. The third considerable limitation is the use of self-reporting scales rather than clinical verification. However, the anonymous nature of the survey and widespread social distancing measures preclude such verification. Additionally, it is not possible to adjust for the confounding effect of non-COVID-19-related individual crisis situations on participant responses. We tried to reduce this effect by formatting survey questions in such a way that would prompt participants to consider their mental state over the preceding week, rather than current mood. Another consideration is limited responses from the WHO African region AFRO, which necessitated exclusion of this very important region from the analysis. It is our hope that similar studies will be conducted in Africa in the future. Finally, a longitudinal assessment of the evolution of psychological symptoms in response to COVID-19 pandemic is imperative and indeed the subject of an ongoing investigation by our group.

Utilizing the ‘feedback’ feature in our online questionnaire, several participants expressed that participation in the survey helped them focus on their mental health. Furthermore, a number of participants reported eating more than usual for comfort or out of boredom. This feedback could aid in efforts to develop mental health screens specific to COVID-19 pandemic.^52^

In conclusion, this effort highlights a significant impact of the COVID-19 pandemic at a regional and worldwide level on the mental health of individuals and elucidates prominent associations with their demographics, history of psychiatric disease risk factors, house-hold conditions, personality traits, and attitude towards COVID-19. These results could serve to inform health professionals and policymakers across the globe, aiding in dynamic optimization of mental health services during and following the COVID-19 pandemic, and reducing its long-term morbidity and mortality.

## Data Availability

All data presented in the main and supplementary items are deposited on the repository below and are available for verification upon request.

https://osf.io/3vupe/?view_only=80f71b6f0c8d49b08573ea12eab10d33

## Acknowledgments

We gratefully acknowledge the contribution of Luciana Armengol (Argentina), Prof. Anthony Hannan, Maxine Mason, Qi Hui Poh (Australia), Taria Brkić (Bosnia and Herzegovina), Barbara Levinsky (Brazil), Alexandra Schimmel and Lea-Caya Bissonnette (Canada), Claudia Valenzuela Rios (Chile), Marc Scherlinger, Alice Tondre, Lola Kouroma, and Morgane Roth (France), Katharina Schlerka, Lisa Garrelts, and Romy Seifert (Germany), Lena Heck (Germany/ Switzerland), Varsha Hooda, Deepak Tanwar, and Chakradhar Yakkala (India), Prof. Mohammad Es haghi and Sepehr Namirad (Iran), Darren Kelly (Ireland), Nour Mosawy (Jordan), Dayra Lorenzo (Mexico), Chirine Katrib (Lebanon/ France), Usman Mukhtar, Uzair Jaswal, and Mubaris Bashir (Pakistan), Prof. Kornelia Kedziora-Kornatowska, Milena Czarnocka and Juli Davis (Poland), Ana Alexandra Moraru (Romania), Shoaib Jawaid (Saudi Arabia/ United Arab Emirates), Myriam Merarchi (Singapore/ France), Michelle McLuckie, Doman Obrist, Niharika Gaur and Graciela Huber (Switzerland), Aurelia Muller (Taiwan/ Germany/ Switzerland), Burak Ozan (Turkey), Carmen Neagoe and Aleena Malik (UK), Anastasiia Timmer (Netherlands/ USA), Colette Rausch, Prof. Paul Schulz, Prof. Mo Salman, Saleha Tahir, Laura Luebbert, Sarish Khan, Rebecca Sager, Lupita Lozano, and American Physician Scientist Association (USA) for their dedicated help in data collection. We are also thankful to Lena Heck and Giuseppe Parente (University of Zurich) for technical support. Finally, we would like to express our gratitude to Prof. Selmira Brkić (Faculty of Medicine, University of Tuzla, Bosnia and Herzegovina), Prof. Leszek Kaczmarek (Director, Nencki-EMBL Braincity, Warsaw, Poland), University of Zurich Research Office, Zurich Cantonal Ethics Commission, Texas Behavioral Health, and European MD-PhD Association for their expedited review of the study procedures under extra-ordinary circumstances and for their organizational support.

## Author contributions

MP and SG contributed in conceptualization, questionnaire development, data collection, data mining, data analysis, visualization, review and editing. RN contributed in data collection, manuscript writing, review and editing. BS, SL, KA, AD, AB, LH, SE, HJ, LRP, VW, BA, MB, and DS contributed in questionnaire translation, data collection, data mining, review, editing, and project co-ordination. PR contributed in data analysis and visualization. ZA contributed in data collection, manuscript writing, review, and editing. ZB contributed in data analysis. ZH and SUQ contributed in data collection and project co-ordination. AMS contributed in data collection, project administration, and editing. AJ contributed in conceptualization, questionnaire development, study approval, data collection, data analysis, data visualization, manuscript writing, review, editing, project administration and supervision. All authors have reviewed and approved the final draft.

## Data and materials availability

https://osf.io/3vupe/?viewonly=80f71b6f=c8d49b08573ea12eab10d33

